# A Step Toward The Use of AI for Polycystic Ovary Syndrome (PCOS): Staged Modelling with Uncertainty-Aware Triage and Conformal Prediction with Cost-Efficient Risk

**DOI:** 10.1101/2025.10.27.25338898

**Authors:** Adewale Alex Adegoke, Idris Babalola, Peter Adebayo Odesola

**Affiliations:** Westminster Foundation for Democracy London, United Kingdom; Department of Health and Social Care London, United Kingdom; Southampton Solent University United Kingdom

**Keywords:** PCOS, staged modelling, cost-aware diagnostics, uncertainty quantification, conformal prediction, clinical utility

## Abstract

Polycystic Ovary Syndrome (PCOS) is a common endocrine disorder affecting women of reproductive age, yet its diagnosis remains challenging due to reliance on costly investigations and limited diagnostic capacity in many clinical settings. We develop and assess a two-stage modelling pipeline in which Stage 1 uses low-cost demographic and routine clinical variables for initial screening, and Stage 2 augments these with laboratory and ultrasound features when escalation is warranted. Logistic Regression (LR) and Random Forest (RF) models are evaluated using calibration and classification metrics (AUC, accuracy, F1, precision, recall, Brier score, ECE), alongside clinical utility via decision curve analysis (DCA). To enhance safety and transparency, we integrate conformal prediction (CP) to provide finite-sample coverage guarantees and controlled abstention. Across train-test performance and out-of-fold evaluations, both models demonstrated consistent performance gains from Stage 1 to Stage 2. AUC increased by 6.9% for LR and increased by 7.4% for RF. LR exhibited more favourable calibration in most settings, while RF achieved higher precision, particularly after escalation. DCA showed higher net benefit for Stage 2 across clinically relevant thresholds. Feature sensitivity analysis indicated that a compact subset of inexpensive predictors preserved over 80% of maximal AUC, supporting cost-efficient screening. CP achieved 94.5% overall coverage with a 41.3% abstention rate, maintaining near-nominal validity across age and BMI subgroups. Capacity-constrained triage experiments showed that prioritising highest-risk cases maximised net benefit when resources were limited. These findings suggest that staged, uncertainty-informed modelling may offer practical steps toward clinically aligned and resource-aware AI support for PCOS assessment.

## Introduction

### 1.1 Background & Motivation

Polycystic ovary syndrome (PCOS) stands as the most common endocrine disorder affecting reproductive-aged women worldwide, with prevalence estimates ranging from 4% to 21% in earlier studies, depending on diagnostic criteria used [1] (Lizneva et al. 2016). Later investigations have reported higher estimates ranging 5% to 26% [2] (D’Souza Pramila et al 2022). This trend demonstrates the increasing attention of the condition and investigation into the global burden of PCOS. In some regions, more recent figures indicate that 8% to 13% of this population may be affected [3] (Tay et al. 2023). The syndrome is a leading cause of anovulatory infertility, accounting for over 75% of cases related to ovulation disorders [4] (Ghafari et al. 2025). Beyond reproductive health, PCOS is associated with significant metabolic and cardiometabolic risks, including insulin resistance, type 2 diabetes, obesity and cardiovascular disease [5] Long et al. (2022) [6] (Hussein et al. 2024). These comorbidities threaten long-term individual health and place considerable socioeconomic burden on healthcare systems. Additionally, delayed diagnosis often as a result of heterogeneous symptom profiles and limitations of conventional diagnostic criteria restrains early intervention strategies aimed at mitigating long-term outcomes [2] (D’Souza Pramila et al 2022). Accurate and timely risk stratification in PCOS is therefore paramount. An effective predictive framework that can swiftly identify high-risk individuals presents a critical opportunity for clinical intervention, resource optimisation and improved patient outcomes.

### 1.2 Gaps in research

While machine learning (ML) applications for PCOS risk prediction are growing in number, they often focus primarily on maximizing diagnostic accuracy using standard performance metrics such as AUC, precision and recall without assessing whether these models deliver meaningful clinical value. A systematic overview noted high reported performance in PCOS detection (AUCs between 73% and 100%, diagnostic accuracy up to 100% and positive predictive value (PPV)/negative predictive value (NPV) frequently exceeding 90%) [7] (Barrera et al, 2023). However, these studies typically lack evaluations based on decision-analytic frameworks that reflect real clinical trade-offs. Moreover, existing work often neglects probabilistic calibration, critical when translating predictions into actionable risk thresholds. Methods to handle predictive uncertainty remain largely unexplored in PCOS contexts, although conformal prediction has shown promise for delivering reliable set-valued predictions in healthcare [8] Vazquez J &, Facelli JC. Likewise, decision curve analysis (DCA) which allows quantification of net clinical benefit across threshold strategies is notably absent from PCOS-focused ML studies [9] (Xu X. et al 2024).

Another important but under-addressed area is operational realism. In many clinical settings, capacity constraints limit how many patients can receive intensive evaluation or screening. Yet, prior PCOS prediction models rarely consider triage strategies under capacity limitations, which can significantly influence utility and practicality. There is limited emphasis on feature cost-efficiency; few studies assess whether omitting low-importance features compromises performance, a key consideration for low-resource implementation. Recent PCOS prediction with machine learning has progressed across three data modalities: self-reported/clinical features, electronic health records (EHRs) and imaging; yet most studies emphasize discrimination (accuracy/AUC) over calibration, decision-analytic value, or uncertainty aware deployment. In a large EHR study, [10] (Zad et al 2024). trained gradient boosting, logistic regression, SVM and random forests on 30,601 at-risk outpatients and reported AUCs of 0.87-0.91 for prevalent PCOS and 0.82-0.84 for incident PCOS, highlighting scalability to routine care but without decision curve analysis (DCA) or coverage-guaranteed uncertainty estimates.

On imaging, [11] (Suha et al 2022). built a deep-learning pipeline on ovarian ultrasound features and achieved 99.89% accuracy in a 1,000-image benchmark, demonstrating near-ceiling discrimination while leaving calibration and clinical utility assessment unexplored. A recent multi-modality study [12] (Abdelsalam, S.H. et al 2024) combined ultrasound with selected clinical predictors and reported 98.29% validation accuracy, again focusing on accuracy gains rather than net clinical benefit or subgroup reliability. Another work by [13] (Agirsoy and Oehlschlaeger 2025) pursued “non-invasive” diagnosis from clinical plus ultrasound features, with XGBoost reaching AUC = 0.995 (Clinical+USG+AMH) and even perfect test-set performance (AUC = 1.00) on a small external set, which the authors themselves caution may reflect overfitting/data leakage, yet calibration, DCA and abstention policies were not explored.

Furthermore, several works develop patient-facing or clinician-support tools from non-invasive or questionnaire-like variables. [14] (Zigarelli et al 2022). proposed “machine-aided self-diagnostic” PCOS models from anthropometrics and symptoms to enable pre-consultation screening; they prioritized convenience and explainability but did not report probability calibration, DCA, or abstention policies. [15] (Shanmugavadivel et al 2024). used correlation-based feature selection to reduce inputs and trained LR/NB/SVM, the SVM achieved 94.44% accuracy on the clinical subset, while a VGG16 modelling pipeline on ultrasound achieved 98.29% validation accuracy again without calibration or decision-analytic evaluation. [16] (Tong et al 2025). built hormone-based diagnostic models combining AMH and steroid/estrogen panels, the logistic classifier achieved test-set ROC-AUC = 0.934 (specificity = 0.947; F1 = 0.864), while XGBoost maximized accuracy (0.905) and sensitivity (0.870), yet no calibration checks, decision curve analysis, or uncertainty aware triage were reported.

A substantial stream of studies relies on the widely used 541-patient Kerala clinical dataset [17] (the same open dataset used in our study), typically targeting binary PCOS classification with conventional metrics. [18] (Khanna et al 2023). introduced an “explainable multi-stacking” framework and reported ~98% accuracy using feature-selected clinical predictors and SHAP-based explanations, but did not assess calibration, capacity-constrained triage, or uncertainty aware abstention. [19] (Rahman et al 2024). built a web-based pipeline on the same dataset, where AdaBoost and Random Forest each reached 94% accuracy with F1 up to 0.91; despite a clear engineering contribution, calibration analysis and DCA were not performed. A comparative study by [20] (Taha et al 2025). evaluated seven supervised learners on the same Kerala PCOS dataset and reported Logistic Regression as best overall (accuracy = 91.7%, precision = 96.0%, ROC-AUC = 96.8%), but it did not assess calibration, decision-analytic utility, uncertainty, or subgroup reliability leaving open questions central to staged, cost-aware deployment and safe abstention. [21] Tiwari et al 2022. (“SPOSDS”) optimized feature selection and classifiers on the same dataset and reported ~93.25% accuracy yet omitted probability calibration diagnostics and decision-analytic evaluation.

In addition, [22] (Danaei Mehr and Polat 2022) systematically compared traditional and ensemble classifiers on the Kerala dataset, underscoring the value of feature selection but again without calibration reporting or uncertainty aware triage. [23] (Bharati et al 2020). previously evaluated logistic regression and random forests on the dataset in IEEE TENSYMP, reporting ~91% accuracy but no assessment of reliability (e.g., calibration curves, Brier score) or clinical net benefit. [24] (Mohi Uddin et al 2025). examined eight traditional and ensemble models on the 541-record dataset, highlighting very high headline scores (e.g., up to 99.78% for LR/SVM in their web supplement) without external validation, calibration, decision-curve, or uncertainty aware triage, highlighting overfitting risks for clinical translation. [25] (Panjwani et al 2025). proposed an optimized stacked ensemble (WaOEL) for “early, inexpensive” PCOS screening from symptomatic, attaining accuracy = 92.8% and AUC = 0.93 on a blended low-cost feature set that included the Kaggle PCOS resource; however, the study emphasized accuracy without calibration, capacity-constrained triage, or formal error-rate control. [26] (Emara et al 2025). introduced a stacked-learning pipeline with ADASYN/SMOTE rebalancing on the same Kaggle dataset and reported 97% accuracy, but again did not quantify probability calibration, net benefit under operational caps, or coverage guarantees. Overall, prior work rarely evaluates models under capacity constraints (e.g., fixed slots for further testing), seldom implements triage policies that integrate model confidence and almost never use formal uncertainty quantification with coverage guarantees in PCOS.

Conformal prediction (CP) has emerged in clinical AI as a distribution-free framework to deliver set-valued predictions with finite-sample coverage, enabling principled abstention; however, CP has not been incorporated into PCOS risk tools. A field primer by [27] (Angelopoulos and Bates 2022) introduced practical CP methods with coverage guarantees but did not target PCOS specifically. A clinical review [8] (Vazquez J, Facelli JC 2022) catalogued CP applications across medical areas and argued for CP to quantify per-case reliability, yet no PCOS application was identified.

Methodological elements needed to bridge research to practice such as DCA for net benefit and calibration for trustworthy probabilities also remain underused in PCOS modeling. DCA is a standard way to quantify whether model-guided action improves decisions over “treat all” or “treat none [28] (Vickers, A.J. & Elkin, E.B 2006) but PCOS studies rarely report it. Even where discrimination is strong (e.g., ultrasound or stacked clinical models), miscalibration can undermine threshold-based actionability [7] (Barrera et al, 2023), a concern not addressed in the above works.

In summary, existing PCOS prediction studies report strong discrimination and excel in accuracy across datasets and modalities but generally (i) do not adopt staged, cost-aware modeling that escalates from low-cost features to richer diagnostics with explicit calibration targets; (ii) do not compare uncertainty aware triage against risk-only or utility-only rankings under capacity limits; and (iii) do not embed conformal predictors to maintain coverage with controlled abstention across subgroups. These are the main motivation for our integrated framework for PCOS risk stratification. Our work is positioned to fill these gaps by integrating staged modeling, decision-analytic evaluation, capacity-aware triage using confidence and conformal prediction into a single deployable pipeline.

### 1.3 Methodological Advances Needed

To address these identified gaps, an improved methodology must combine clinical decision-theoretic evaluation, calibration, uncertainty quantification and cost-aware feature selection in a coherent workflow. Models should be assessed for accuracy alongside clinical net benefit, using decision curve analysis (DCA). DCA helps determine whether a model-based decision is better than treating all or none, at relevant threshold probabilities and has been particularly influential in evaluating diagnostic tools [28] (Vickers, A.J. & Elkin 2006). Ensuring that predicted probabilities are well-calibrated is critical, metrics such as the Brier score and expected calibration error (ECE) help measure this. In healthcare, even well-performing classifiers can be misleading if miscalibrated, leading to harmful clinical decisions [29] (Van Calster et al., 2019).

Furthermore, embracing uncertainty aware prediction frameworks, such as conformal prediction, offers a structured way to generate prediction sets with finite-sample coverage guarantees, allowing the system to abstain when confidence is low. This approach has gained traction for its ability to improve trustworthiness and fairness in clinical AI [27] (Angelopoulos & Bates, 2022). Triage or selection policies under capacity constraints must be modelled explicitly because real-world healthcare settings often cannot screen or follow up with all patients. Developing risk and utility-based triage protocols under fixed caps ensures models are evaluated in a realistic operational context. In addition, assessing feature cost-efficiency via feature importance and drop-sensitivity analysis helps identify minimal input sets that maintain performance. This is essential when deploying AI tools in settings where data collection is expensive or limited, such as low-resource clinics.

### 1.4 Research Question

This study is guided by three core research questions that together address performance, operational feasibility and safety in AI-assisted PCOS risk prediction:

1. How can a staged AI modelling framework, starting with low-cost clinical features and escalating to richer diagnostics, support potential improvements in predictive accuracy for PCOS and calibration in clinical risk prediction while maintaining cost-efficiency?
2. Under real-world capacity constraints, to what extent can uncertainty aware triage strategies offer advantage over conventional risk-based and utility-based prioritisation in term of net clinical benefit?
3. How can conformal prediction maintain high coverage and controlled abstention rates across PCOS patient subgroups and how can this contribute to safer and trustworthy AI-assisted decision-making?

These questions form the backbone of our investigation, linking model development with downstream clinical decision support, safety considerations and responsible use.

### 1.5 Contribution and Novelty of This Work

This study presents a proof-of-concept, unified framework for AI assisted PCOS risk stratification that integrates performance evaluation, cost considerations, uncertainty handling and clinical utility. The framework is built around four components that jointly assess performance, interpretability, resource constraints and safety rather than in isolation.

We summarize the contributions of our work as follows:

1. By building models in a two-stage process starting with inexpensive clinical features and optionally enriching them with additional low-cost predictors, we demonstrate that substantial performance gains both in discrimination and calibration can be achieved. Feature importance and sensitivity analysis revealed that retaining a subset of carefully selected features preserved upwards of 80% of maximal AUC performance, a strategy in line with efforts in clinical ML to maintain accuracy while reducing data collection burden [30] (Moons et al., 2015) [31] (Steyerberg, 2019).
2. We extend model evaluation beyond conventional accuracy metrics by incorporating Decision Curve Analysis (DCA) to quantify clinical net benefit across plausible decision thresholds. This allows model performance to be assessed in terms of its potential impact on clinical decision-making, an approach increasingly advocated in prognostic research but still limited in PCOS-focused applications [32] (Moons et al., 2009).
3. We operationalise uncertainty aware triage policies, integrating model confidence with risk scores to make smarter prioritisation decisions under capacity constraints. Unlike standard risk-ranking, this approach optimises net benefit while avoiding over or under-treatment, a concept emerging in triage-supporting AI systems but rarely tested in PCOS contexts.
4. We embed conformal prediction within the modelling pipeline to produce set-valued predictions with finite-sample coverage guarantees and explicit abstention. This enables systematic identification of high-uncertainty cases for escalation or review and supports more equitable performance across subgroups such as age and BMI, contributing to safer and more transparent AI-assisted decision support [33] (Olsson et al., 2022).

The following section describes the data, modelling approaches, evaluation metrics and decision-analytic methods used to address these questions.

## 2 Methods

### 2.1 Data Description

The dataset used in this study was obtained from the publicly available Polycystic Ovary Syndrome (PCOS) repository on Kaggle. The data was originally collected from 10 different hospital across Kerala, India. It contains 541 patient records and 44 variables, including a binary target variable indicating the presence of PCOS (1) having 177 participants (32.7%) or absence (0) of PCOS having 374 participants (67.3%). The dataset does not provide verifiable diagnostic criteria for the PCOS label. The variables capture demographic information, anthropometric measurements, lifestyle habits, menstrual history, biochemical assay results and ultrasound findings.

### 2.2 Data Pre-processing

All column names were stripped of extra spaces to ensure consistency. A missing value check identified two variables with incomplete records:

- Marriage Status (Years) - 1 missing value, handled using median imputation.
- Fast food (Y/N) - 1 missing value, imputed with the mode.

We selected these imputation methods due to the minimal proportion of missing data and their ability to preserve the central tendency of the variables with minimal distortion. [34] *Zhang Z. 2016*

Duplicate record checks across all variables returned zero duplicates. Binary categorical variables such as Pregnant(Y/N), Weight gain(Y/N) were identified by the (Y/N) suffix in their names and recorded as integers (0 = No, 1 = Yes) to facilitate direct use in analysis. After these steps, the dataset contained 541 complete observations and 44 variables, with no missing values or duplicates.

### 2.3 Feature Partitioning

For modelling purposes, we separated all predictor variables into two clinically motivated categories based on the method and cost of acquisition. The target variable (PCOS (Y/N)) is a binary indicator of confirmed diagnosis. In clinical practice, PCOS diagnosis often follows a staged process:

- Initial screening using low-cost, easily obtainable information such as patient history, anthropometric measurements and routine vital signs.
- Confirmatory testing using resource-intensive procedures such as hormonal assays and pelvic ultrasound.

Fig 2 below presents an overview of the proposed staged, cost-aware screening framework for PCOS. Low-cost features are used for initial screening, with uncertainty guiding escalation to higher-cost assessment. The workflow emphasises efficient resource use and transparent decision support rather than algorithmic complexity.

**Fig 1:**
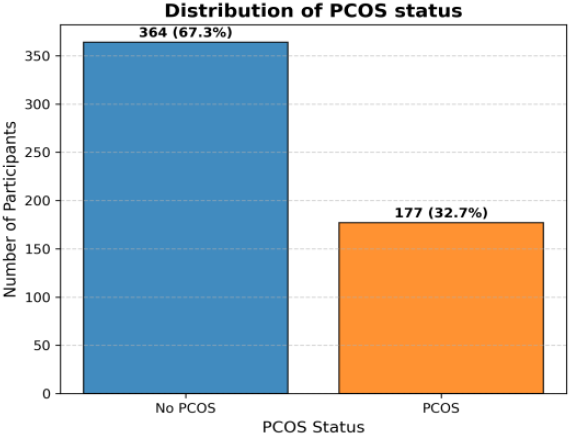
Distribution of PCOS Status.

**Fig 2:**
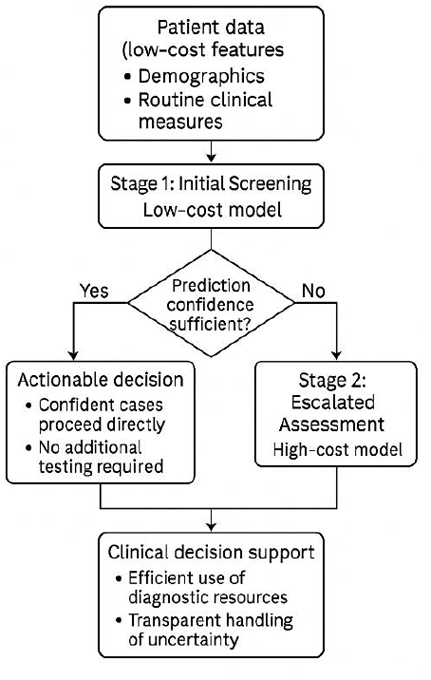
Proposed workflow of the staged PCOS modelling.

The dataset was partitioned to reflect this workflow, enabling evaluation of staged diagnostic pathways that mirror real-world resource constraints. Table 2 captures the criteria used for the selection of both inexpensive and expensive features for the staged modelling. We present the resultant feature partitioning in Table 3.

**Table 1:** Variables with their data types and brief descriptions.

| Feature Name | Data Type | Description |
| --- | --- | --- |
| Age (years) | Numeric | Age of the patient in years |
| Weight (Kg) | Numeric | Body weight measured in kilograms |
| Height (Cm) | Numeric | Height of the patient in centimetres |
| BMI | Numeric | Body mass index, calculated from weight and height |
| Blood Group | Categorical | Patient's blood group, encoded numerically |
| Pulse rate(bpm) | Numeric | Heart rate measured in beats per minute |
| RR (breaths/min) | Numeric | Respiratory rate measured in breaths per minute |
| Hb(g/dl) | Numeric | Haemoglobin concentration in grams per decilitre |
| Cycle(R/I) | Categorical | Menstrual cycle regularity or irregularity indicator |
| Cycle length(days) | Numeric | Length of menstrual cycle in days |
| Marriage Status (Years) | Numeric | Number of years since marriage |
| Pregnant(Y/N) | Binary | Pregnancy status |
| No. of abortions | Numeric | Number of abortions or miscarriages |
| I beta-HCG (mIU/mL) | Numeric | Serum beta-HCG hormone level (first measurement) |
| II beta-HCG (mIU/mL) | Numeric | Serum beta-HCG hormone level (second measurement) |
| FSH (mIU/mL) | Numeric | Follicle-stimulating hormone level |
| LH (mIU/mL) | Numeric | Luteinising hormone level |
| FSH/LH | Numeric | Ratio of FSH to LH |
| Hip(inch) | Numeric | Hip circumference in inches |
| Waist(inch) | Numeric | Waist circumference in inches |
| Waist:Hip Ratio | Numeric | Waist-to-hip circumference ratio |
| TSH (mIU/L) | Numeric | Thyroid-stimulating hormone level |
| AMH (ng/mL) | Numeric | Anti-Müllerian hormone level |
| PRL (ng/mL) | Numeric | Prolactin hormone level |
| Vit D3 (ng/mL) | Numeric | Vitamin D3 level |
| PRG (ng/mL) | Numeric | Progesterone hormone level |
| RBS (mg/dl) | Numeric | Random blood sugar level |
| Weight gain(Y/N) | Binary | Indicator for recent weight gain |
| hair growth(Y/N) | Binary | Indicator for excessive hair growth |
| Skin darkening (Y/N) | Binary | Indicator for skin hyperpigmentation |
| Hair loss(Y/N) | Binary | Indicator for hair loss |
| Pimples(Y/N) | Binary | Indicator for presence of acne |
| Fast food (Y/N) | Binary | Indicator for regular fast-food consumption |
| Reg.Exercise (Y/N) | Binary | Indicator for regular exercise |
| BP Systolic (mmHg) | Numeric | Systolic blood pressure measurement |
| BP Diastolic (mmHg) | Numeric | Diastolic blood pressure measurement |
| Follicle No. (L) | Numeric | Number of ovarian follicles on the left ovary |
| Follicle No. (R) | Numeric | Number of ovarian follicles on the right ovary |
| Avg. F size (L) (mm) | Numeric | Average follicle size in the left ovary |
| Avg. F size (R) (mm) | Numeric | Average follicle size in the right ovary |
| Endometrium (mm) | Numeric | Endometrial thickness in millimetres |

**Table 2:** Principles of Feature Partitioning.

| Stage | Inclusion Criteria | Examples |
| --- | --- | --- |
| **Stage 1 - inexpensive features** | Self-reported, physical examination, or routine vital signs; no laboratory or imaging required; minimal acquisition cost. | Age, BMI, Blood Pressure, Lifestyle indicators |
| **Stage 2 - Expensive features** | Laboratory analysis (e.g., hormonal assays) or imaging procedures (e.g., pelvic ultrasound); require specialist facilities and higher financial cost. | FSH, LH, AMH, Ultrasound follicle counts |

**Table 3:**
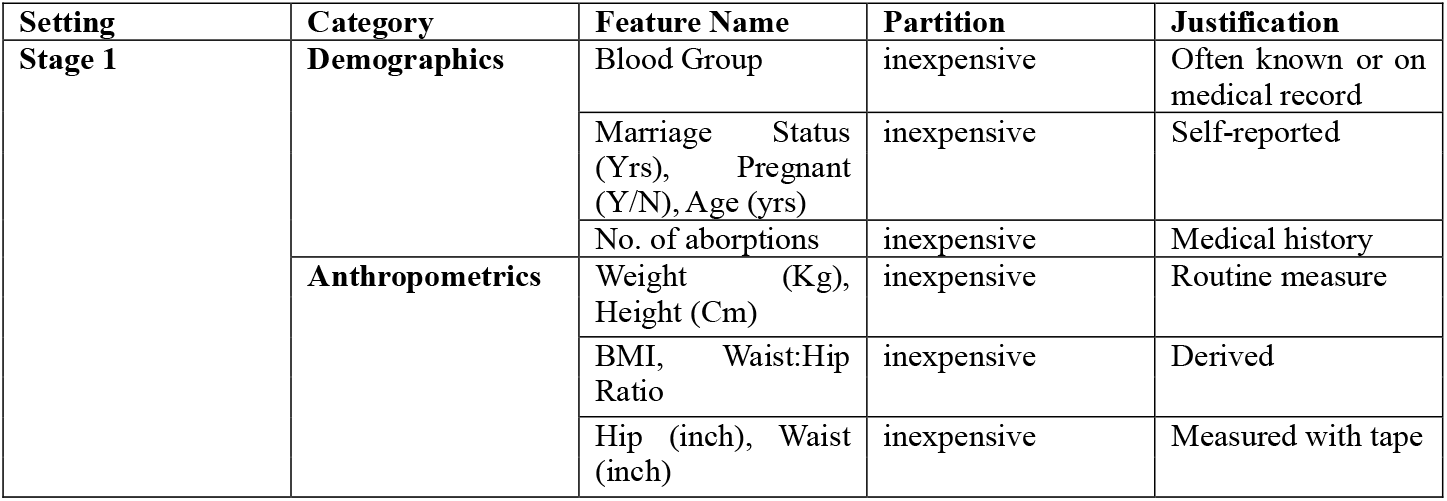
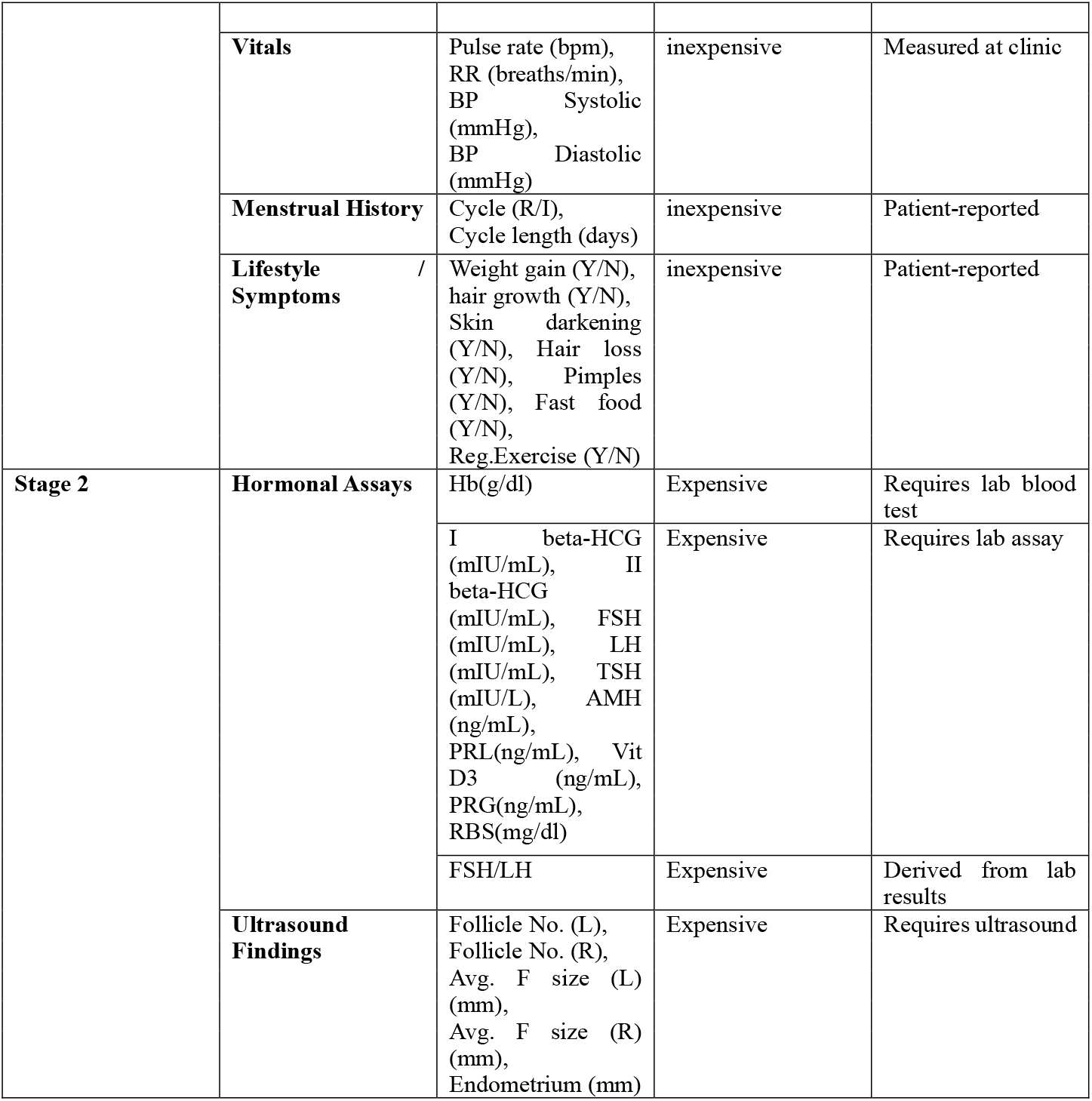
Feature Categories after partitioning with Justification.

The Stage 1 set comprised 24 inexpensive features while the Stage 2 dataset extended Stage 1 by adding predictors that required laboratory analysis (i.e inexpensive + expensive features). This split reflects real-world diagnostic economics; Stage 1 features are low-cost or free to collect, while Stage 2 features require specialized lab or imaging services. This partitioning allows:

1. Stage 1 models trained solely in inexpensive features to serve as screening tools in low-resource settings.
2. Stage 2 models trained on all available features to provide upper-bound diagnostic performance when full testing is feasible.
3. Direct evaluation of decision-focused triage strategies that select a subset of patients for Stage 2 diagnostics based on Stage 1 predictions and uncertainty quantification.

### 2.4 Baseline Modelling Framework

#### 2.4.1 Modelling Stages

The baseline modelling was structured into two stages, aligned with the resource-availability framework introduced in Section 2.3. We developed Stage 1 models using only the “inexpensive” feature set, representing variables obtainable at low cost and minimal clinical intervention, thus simulating a feasible screening process in primary care or community health environments. Stage 2 models incorporated both inexpensive and “expensive” features, enabling evaluation under a fully resourced diagnostic setting. This staged design facilitated direct performance comparisons between minimal and complete resource utilisation, while providing a structured basis for subsequent triage-based decision-making analysis.

#### 2.4.2 Baseline Algorithms

We implemented two supervised classification algorithms for both stages to establish baseline performance benchmarks:

1. Logistic Regression (LR), serving as an interpretable linear model, with standardisation applied to all numeric predictors. Its transparency allows direct inspection of coefficient magnitudes and directions, supporting clinical interpretability [31] (Steyerberg, 2019).
2. Random Forest (RF), an ensemble of decision trees capable of modelling non-linear relationships and higher-order feature interactions, with intrinsic robustness to multicollinearity and noise [35] (Breiman, 2001).

Our aim in this study was to evaluate a staged, uncertainty-aware, cost-sensitive framework rather than to compare a broad set of algorithms. We therefore selected LR and RF because they are well-established, computationally efficient methods that let us test whether the staged approach can deliver meaningful stratification without relying on complex or resource-intensive models. This proof-of-concept focus allowed us to investigate the value of the framework itself while keeping computation lightweight and practical for potential clinical use. The training approaches for this work in summarised in Table 4.

**Table 4:**
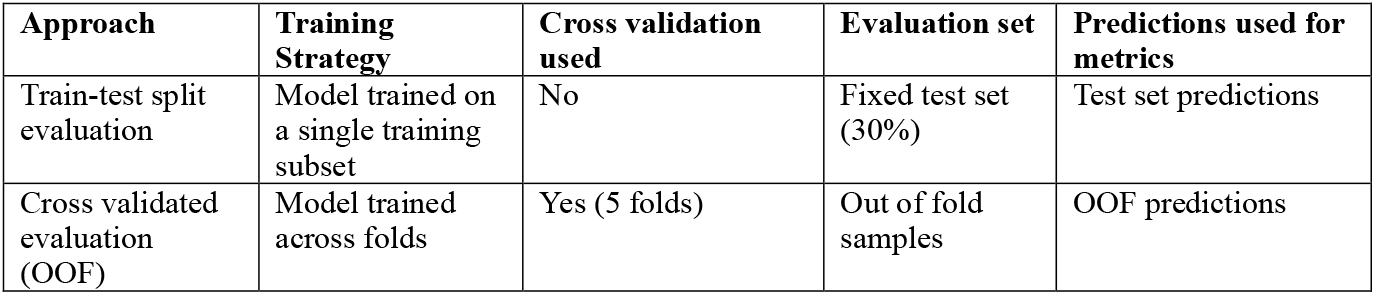
Model training framework.

| Approach | Training Strategy | Cross validation used | Evaluation set | Predictions used for metrics |
| --- | --- | --- | --- | --- |
| Train-test split evaluation | Model trained on a single training subset | No | Fixed test set (30%) | Test set predictions |
| Cross validated evaluation (OOF) | Model trained across folds | Yes (5 folds) | Out of fold samples | OOF predictions |

#### 2.4.3 Evaluation Metrics

We assessed model performance using a combination of statistical and clinical evaluation measures. Discrimination ability was quantified via the Area Under the Receiver Operating Characteristic Curve (AUC), Accuracy, F1-score, Precision and Recall [36] (Powers, 2011). Calibration quality was measured using the Brier score [37] (Brier, 1950) and Expected Calibration Error (ECE) [38] (Naeini et al., 2015), ensuring predicted probabilities aligned with observed event rates. In addition, decision curve analysis [28] (Vickers & Elkin, 2006) was retained in the evaluation plan to estimate clinical net benefit across a range of threshold probabilities, enabling direct translation of model outputs into decision-focused frameworks to be explored in Section 2.5.

#### 2.4.4 Integrated Feature Importance and Selection

In stage 1, permutation importance [39] (Altmann et al., 2010) was applied to quantify the marginal contribution of each inexpensive feature. To evaluate the robustness of these rankings and assess potential for cost-efficient feature reduction, we performed a supplementary bottom-k sensitivity analysis. The K least important inexpensive features (ordered by permutation importance) were removed in sets of K = 3, 5 and 8, and model performance was re-estimated. This analysis allowed us to understand whether removing inexpensive variables could simplify the model while preserving or improving discrimination.

For the expensive feature set in Stage 2, we conducted a one-at-a-time ablation analysis. Each high-cost feature was removed individually and the resulting change in AUC (ΔAUC) was recorded. This directly quantified the marginal utility of each advanced diagnostic measurement, allowing us to identify the features that most substantively improved predictive performance.

The choice of distinct feature-selection approaches reflects the different aims of the two stages and how they work together in a cost-sensitive workflow. Stage 1 aims to identify the minimal set of low-cost features that support early triage, making permutation-based ranking and grouped removal appropriate for evaluating redundancy. Stage 2 requires determining which costly tests meaningfully enhance discrimination beyond Stage 1, for which single-feature ablation provides the clearest estimate of marginal gain. The two analysis therefore offer complementary insights; Stage 1 isolates informative low-cost variables for initial PCOS risk assessment, while Stage 2 identifies high-cost features that justify escalation.

#### 2.4.5 Preventing Data Leakage

All preprocessing, feature selection and model fitting procedures were performed strictly within the training folds during cross-validation, with transformations applied to validation and test folds only after fitting. This ensured no information from the validation or test data influenced model training, thereby preserving the validity of out-of-sample performance estimates [40] (Kaufman et al., 2012).

The outputs from the baseline modelling phase serve three primary functions:

1. Identification of high-yield inexpensive features suitable for low-resource screening (Stage 1).
2. Quantification of expensive feature via ablation and cost-benefit analysis (Stage 2).
3. Provision of benchmark performance metrics for integration into decision curve analysis and conformal prediction frameworks in subsequent sections.

By explicitly quantifying feature contributions, calibration behaviour and performance trade-offs under different resource constraints, the baseline framework provides a robust empirical foundation for the advanced decision-support strategies that follow.

#### 2.4.6 Baseline Model Experimental Settings

To ensure reproducibility, we conducted all experiments with fixed parameters as summarised in Table 5. These parameters detail the data partitioning strategy, cross-validation configuration, calibration settings and algorithm-specific parameters.

**Table 5:** Baseline Model Experimental Settings.

| Parameter | Value |
| --- | --- |
| Train/test split ratio | 70% training / 30% testing (stratified by target) |
| Cross-validation folds (OOF) | 5 folds (StratifiedKFold) |
| Random seed | 42 (fixed across all experiments) |
| ECE bin count | 10 bins |
| Random forest trees (Stage models) | 500 trees |
| Random forest trees (Permutation importance) | 200 trees |
| Scaling method | StandardScaler (applied only to LR numeric features) |
| Evaluation metric for model selection | AUC (ROC area) |

**Table 6:**
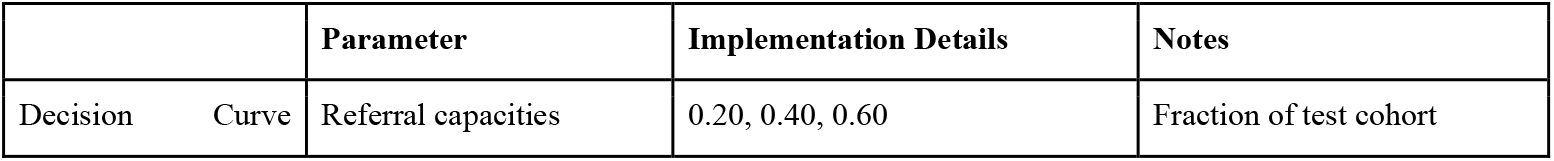
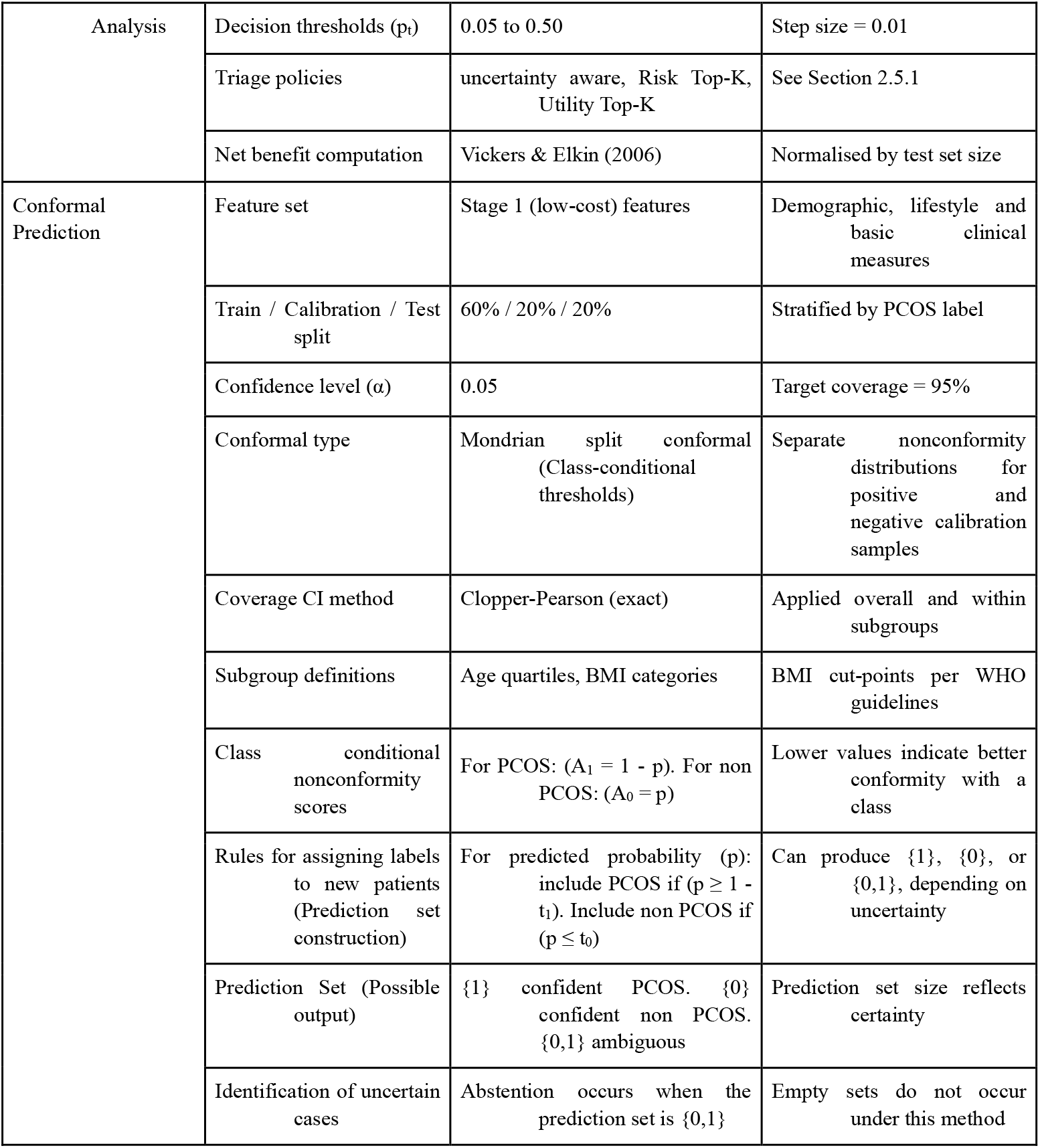
Experimental Settings for decision curve analysis & conformal prediction.

**Table 7:** Experimental Settings for Statistical analysis.

| Setting | Value |
| --- | --- |
| ECE computation | 10 probability bins, bootstrap CIs from 500 resamples |
| Brier score differences | Paired bootstrap with 1,000 resamples |
| Net benefit comparisons | Bootstrap with 2,000 stratified patient-level resamples |
| Conformal coverage evaluation | Nominal confidence level per method (reported in results) |

### 2.5 Decision-Focused Learning with Conformal Prediction

In this phase of the study, we extended our predictive framework by integrating decision-focused learning with class-conditional split conformal prediction [41] (Vovk et al., 2005; [42] Shafer & Vovk, 2008). We optimised downstream clinical decision-making while maintaining formal, distribution-free coverage guarantees. This integration allowed us to embed safety constraints directly into the triage process and to ensure that uncertainty quantification was equitable across patient subgroups.

#### 2.5.1 Clinical Utility Assessment and Capacity-Constrained Referral Optimisation

In this study, we evaluated the clinical utility of the PCOS risk models and their operational use under constrained diagnostic capacity using a decision-analytic framework. Fig 3 illustrates the flow from Stage 1 and Stage 2 PCOS risk modelling through decision curve analysis, net benefit evaluation across thresholds, and comparison of risk-based, utility-based, and uncertainty-aware referral strategies under fixed diagnostic capacity constraints we experiment in this study.

**Fig 3:**
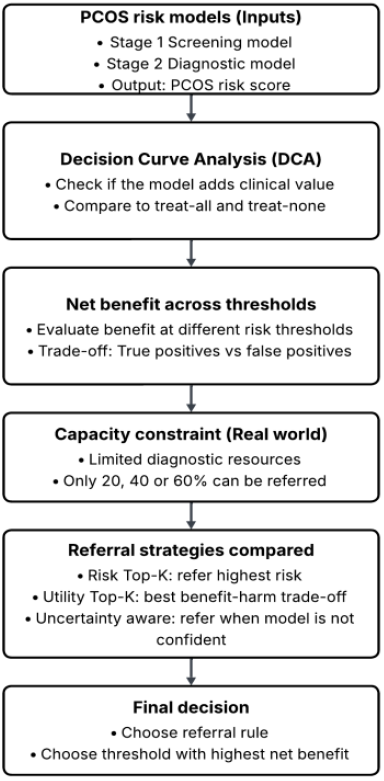
Clinical utility and capacity-constrained decision framework for our PCOS risk models.

We used predicted PCOS risk scores from both the Stage 1 screening model and the Stage 2 diagnostic model as inputs to this process. We first assessed model usefulness using Decision Curve Analysis (DCA) [28], which quantifies clinical value in terms of net benefit across a range of decision thresholds. For each threshold probability, we compared model-based referral decisions against two non-informative reference strategies: treating all individuals as high risk and treating none. Net benefit was computed as

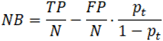

Where TP = True Positive, FP = False Positive, N = patient population, *p*_*t*_ = Decision thresholds

We then extended the analysis to reflect real-world constraints on diagnostic capacity. In this work, we considered fixed referral capacities of 20%, 40%, and 60%, representing scenarios in which only a subset of women can be referred for further assessment. Within each capacity setting and at each decision threshold, we evaluated three referral strategies.

First, we considered a Risk Top-K policy, in which referrals were allocated to individuals with the highest predicted PCOS risk probabilities. This strategy reflects a straightforward risk-ranking approach, in which referral decisions are based solely on estimated disease likelihood, without explicitly accounting for the relative costs of false positive versus false negative decisions at a given threshold. Second, we evaluated a Utility Top-K policy, which ranked individuals according to an expected utility score

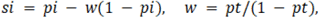

Third, we evaluated an Uncertainty-Aware policy, in which individuals for whom the model was unable to make a confident classification were prioritised for referral, and any remaining referral capacity was subsequently allocated using the same utility-based ranking described above. This strategy is designed to ensure that diagnostically ambiguous cases are not systematically deprioritised under capacity constraints, while still maintaining alignment with threshold-specific decision utility.

For each referral capacity, we computed net benefit across the full threshold grid for all three strategies and identified the threshold that maximised net benefit. This analysis defines the optimal capacity-constrained referral rules for PCOS risk stratification and provides the decision-analytic foundation for the uncertainty-aware triage framework used in this study.

#### 2.5.2 Data Partitioning for Conformal Prediction

We partitioned the dataset into training, calibration and test sets using a stratified split to preserve class balance. In line with established conformal methodology [41] (Vovk et al., 2005; [43] Barber et al., 2021), we allocated 60% of the data for training, 20% for calibration and 20% for testing. We used the calibration set exclusively for determining nonconformity thresholds, thereby ensuring an unbiased evaluation on the held-out test set.

#### 2.5.3 Model Training and Nonconformity Scoring

We trained a classifier [35] (Breiman, 2001) on the training set. Using the calibration set, we obtained class-conditional probability estimates and computed nonconformity scores: for positive cases, (1 - p_1_); for negative cases, (p_1_), where p_1_ denotes the predicted probability of the positive label. We then calculated class-conditional (Mondrian) quantiles using the finite-sample formula:

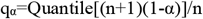

with α = 0.05, yielding separate thresholds t_1_ and t_0_ for the positive and negative classes.

#### 2.5.4 Prediction Set Construction and Subgroup Analysis

For each test instance, we constructed the conformal prediction set according to inclusion rules: we included class 1 if p_1_ >= 1 - t_1_ and class 0 if p_1_ <= t_0_. When both conditions were satisfied, we labelled the case as abstained, triggering a deferred decision. We then evaluated coverage and abstention rates across clinically relevant subgroups, including age quartiles and BMI categories defined by WHO guidelines.

#### 2.4.5 Coverage and Confidence Interval Estimation

We computed coverage, abstention rates, singleton set rates and polarity-specific singleton proportions both overall and within subgroups. To quantify uncertainty in these estimates, we applied the Clopper-Pearson exact method [44] (Clopper & Pearson, 1934), which provided reliable confidence intervals even for smaller subgroup sizes.

### 2.6 Comparative Evaluation and Statistical Analysis

#### 2.6.1 Evaluation Strategy

We designed a unified evaluation framework to compare baseline classifiers, decision-focused learning models and their conformal prediction-augmented variants. Our analysis jointly assessed predictive performance, calibration reliability, clinical decision utility and uncertainty quantification, with all statistical inference grounded in out-of-fold (OOF) predictions. These OOF predictions served as the common input for all subsequent statistical tests, calibration analysis, decision curve estimations and bootstrap confidence interval computations.

#### 2.6.2 Performance, Calibration and Decision Utility Analysis

We evaluated discrimination using the area under the ROC curve (AUC), with statistical differences between paired AUCs tested via the DeLong method for correlated ROC curves [45] (DeLong et al., 1988). Sensitivity and specificity were compared using the McNemar test for paired binary classifications, applied at a fixed decision threshold of 0.5. Calibration was assessed through both the Brier score and the expected calibration error (ECE), the latter computed with 10 equal-width probability bins. ECE estimates were accompanied by bootstrap-derived 95 % confidence intervals using 500 stratified resamples to preserve class proportions. Paired differences in Brier scores between models were quantified using bootstrap mean difference estimation (1,000 resamples).

For uncertainty quantification, we examined empirical coverage rates from the conformal prediction sets both overall and within clinically relevant strata (PCOS-positive vs. negative, age quartiles and BMI categories). Deviations from nominal coverage were tested using exact binomial confidence intervals (Clopper-Pearson) and abstention rates were compared between subgroups via the Chi-square test. To assess clinical utility, we applied decision curve analysis [28] (Vickers & Elkin, 2006) and simulated diagnostic capacity constraints allowing advanced testing for 20 %, 40 % and 60 % of patients. For each scenario, we computed net benefit (NB) and the number of unnecessary advanced tests avoided. Pairwise NB differences were evaluated via bootstrap confidence intervals using 2,000 stratified patient-level resamples.

#### 2.6.3 Statistical Inference and Multiple Comparisons

All hypothesis tests were two-sided, and p-values were adjusted for multiple comparisons using the Benjamini-Hochberg false discovery rate (FDR) control procedure. Statistical significance was defined as an adjusted p < 0.05. Effect sizes and CIs were prioritised over raw p-values in interpretation to align with modern statistical reporting recommendations [46] (Wasserstein et al., 2019).

Bootstrapping, exact tests and non-parametric alternatives (Wilcoxon signed-rank) were applied when distributional assumptions could not be verified via the Shapiro-Wilk test. All analysis were implemented in Python using NumPy, pandas and custom-written functions for DeLong, McNemar and bootstrap procedures, ensuring complete reproducibility without reliance on opaque third-party wrappers.

## 3. Results

### 3.1 Baseline Modelling Performance

In this study we evaluated both the train-test performance (baseline) and the out-of-fold (OOF) cross-validation estimates for Stage 1 (inexpensive-only) and Stage 2 (inexpensive + expensive) models, reporting discrimination (AUC, %), Accuracy (%), F1-score (%), Precision (%), Recall (%) and pre-calibration metrics (Brier score and ECE) summarised in Table 8, following TRIPOD guidelines for transparent reporting of prediction models [47] Collins et al., 2015.

**Table 8:**
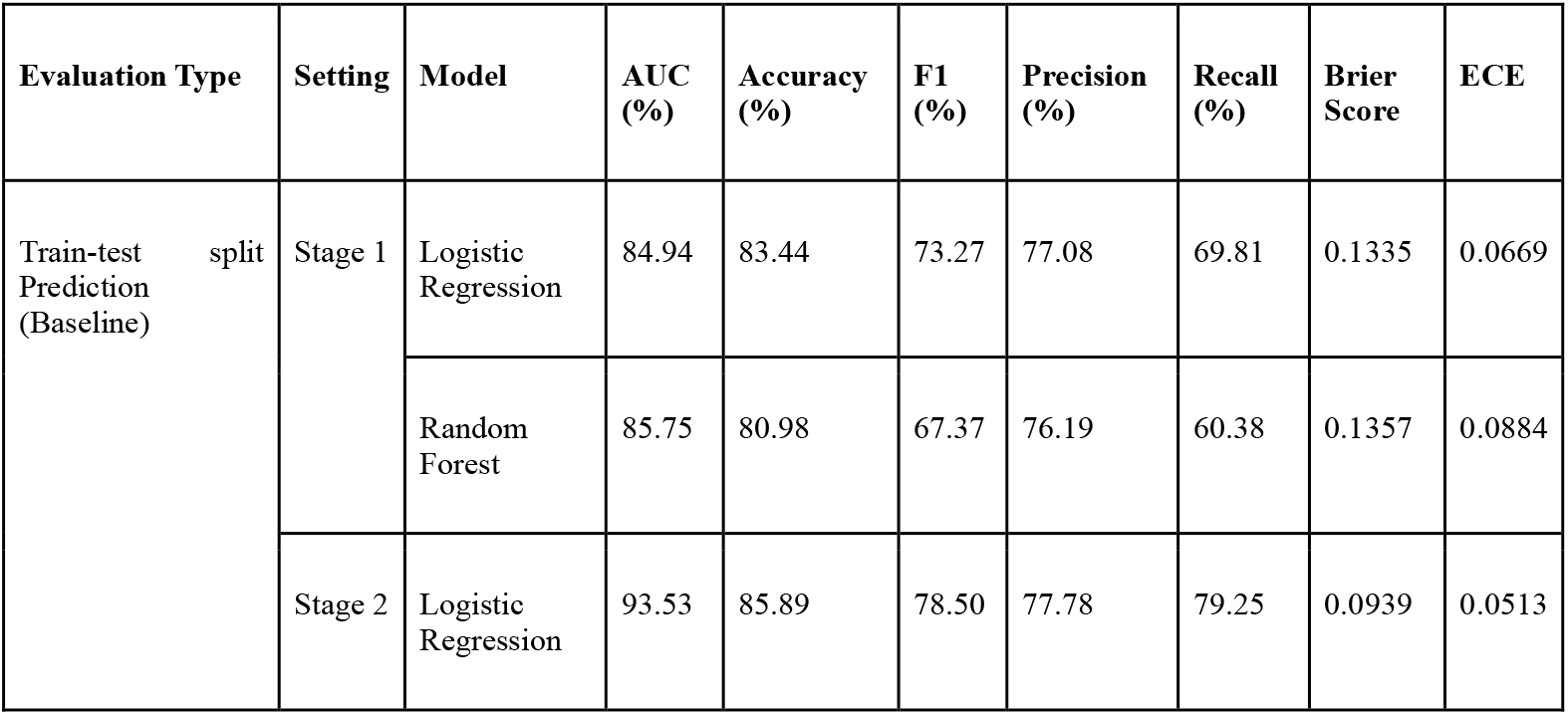
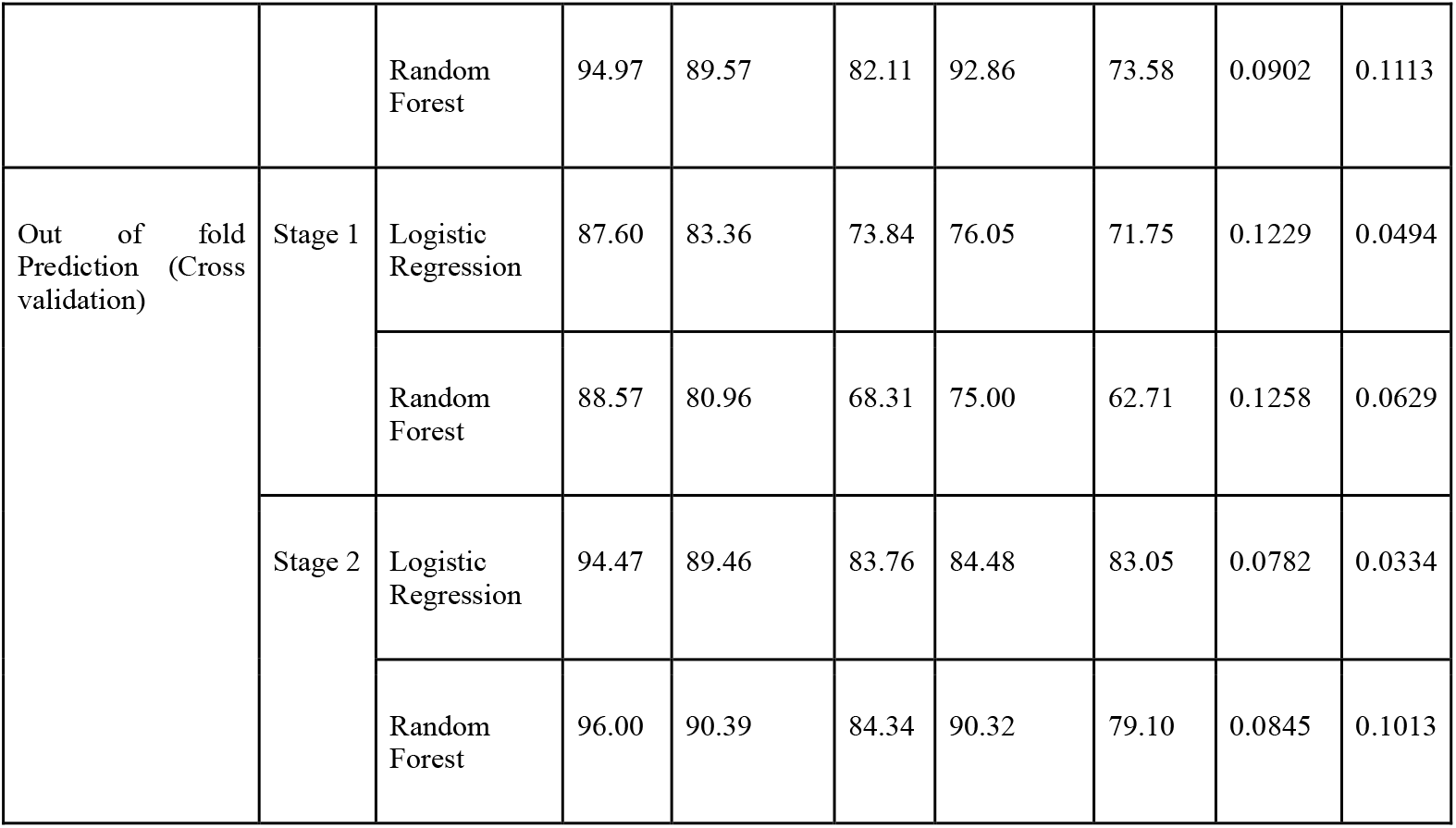
Train-test split performance and out-of-fold performance metrics (AUC, F1, Precision, Recall, Brier, ECE) for Stage-1 (inexpensive) and Stage-2 (inexpensive + expensive) Logistic Regression and Random Forest model.

For Stage 1, the train-test split results showed broadly comparable performance for Logistic Regression (LR) and Random Forest (RF): LR achieved 84.94% AUC, 83.44% Accuracy, 73.27% F1, 77.08% Precision and 69.81% Recall with Brier = 0.1335 and ECE = 0.0669, while RF reached 85.75% AUC, 80.98% Accuracy, 67.37% F1, 76.19% Precision and 60.38% Recall with Brier = 0.1357 and ECE = 0.0884. The corresponding OOF estimates were slightly higher for discrimination and accuracy and exhibited better pre-calibration, with LR at 87.60% AUC, 83.36% Accuracy, 73.84% F1, 76.05% Precision, 71.75% Recall, Brier = 0.1229, ECE = 0.0494 and RF at 88.57% AUC, 80.96% Accuracy, 68.31% F1, 75.00% Precision, 62.71% Recall, Brier = 0.1258, ECE = 0.0629 as presented in Table 8. When we experimented with inexpensive-only features, LR provided a more balanced sensitivity-specificity profile and more favorable pre-calibration than RF, which tended to prioritise precision over recall.

When moving to Stage 2, where advanced laboratory and ultrasound features are added, for both algorithms in both evaluations. On the train-test approach, LR obtained 93.53% AUC, 85.89% Accuracy, 78.50% F1, 77.78% Precision, 79.25% Recall, with Brier = 0.0939 and ECE = 0.0513, while RF achieved 94.97% AUC, 89.57% Accuracy, 82.11% F1, 92.86% Precision, 73.58% Recall, Brier = 0.0902, ECE = 0.1113. The OOF estimates were again higher overall and showed the same qualitative pattern: LR reached 94.47% AUC, 89.46% Accuracy, 83.76% F1, 84.48% Precision, 83.05% Recall, with Brier = 0.0782 and ECE = 0.0334; RF achieved 96.00% AUC, 90.39% Accuracy, 84.34% F1, 90.32% Precision, 79.10% Recall, with Brier = 0.0845 and ECE = 0.1013. We evaluated these pre-calibrated ECE and Brier values to characterise raw probability quality; LR exhibited lower ECE than RF, indicating less over-confidence, whereas RF delivered higher precision with a moderate penalty in calibration.

These results illustrate the expected pattern in a staged diagnostic framework where Stage 1 models provide moderately high discrimination with minimal cost, while Stage 2 models benefiting from specialised tests deliver near optimal classification performance to enhance better decision making in clinical triage and resource allocation. The baseline results align with the cross-validated OOF estimates: Stage 2 (inexpensive + expensive) materially improves discrimination and for LR, pre-calibration relative to Stage 1 (inexpensive-only). Between models, RF offers the strongest precision especially in Stage 2 while LR yields more balanced recall and better raw probability calibration. These findings support a staged diagnostic workflow in which Stage 1 enables cost-efficient screening and Stage 2 provides superior accuracy once confirmatory tests are available.

Next, we plot the ROC curves used to compare the two-stage modelling pipeline for the OOF estimates: Stage 1 and Stage 2 in Fig 4. ROC curves evaluate the trade-off between sensitivity (true positive rate) and specificity (false positive rate) across varying decision thresholds [48] (Fawcett, 2006). In Stage 1, the Random Forest (RF) achieved an AUC of 88.60%, slightly outperforming Logistic Regression (LR) with an AUC of 87.6%. This indicates both models have strong discriminative power, but RF captures marginally more non-linear interactions in the limited feature set In Stage 2, incorporating expensive diagnostic features alongside the inexpensive set, both models improved markedly. RF rose to an AUC of 96.00%, while LR increased to 94.47%. The improvement is consistent with prior findings that richer feature sets enhance classification performance, particularly for ensemble methods [49] (Noroozi, Z et al 2023). The steeper rise of the ROC curve near the y-axis in Stage 2 reflects a reduction in false positives at high sensitivity levels. While LR’s AUC gains from Stage 1 to Stage 2 were substantial, RF retained a consistent advantage at both stages. This aligns with the known robustness of tree-based ensembles to complex feature interactions and heterogeneous data scales [50] (Cutler, D. R et al 2007) [51] (García-Carretero, R et al 2021), suggesting RF may be better suited when combining heterogeneous “inexpensive” and “expensive” features in a hybrid screening pipeline.

**Fig 4:**
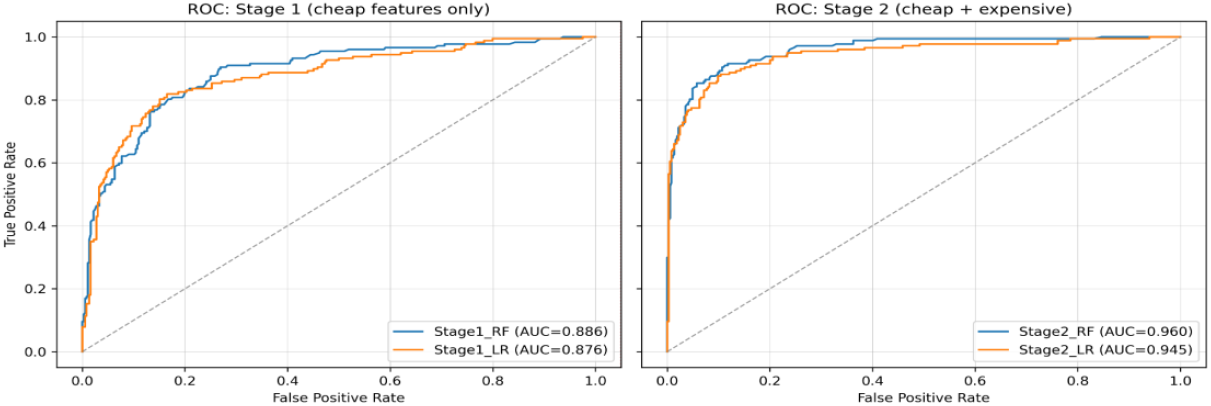
ROC curves for Stage 1 and Stage 2 models. Curves plotted from OOF predictions.

We also assessed how well predicted probabilities matched observed outcomes using reliability diagrams built from OOF predictions shown in Fig 5. Probabilities were partitioned into 10 equal-width bins and each point was annotated with its bin count to make sampling variability explicit. In Stage 1 (inexpensive features only), both Random Forest and Logistic Regression track the diagonal closely across the bulk of the probability range. Small departures appear around the mid-probability bins (≈0.3-0.6), but the curves remain near the ideal line especially for Stage 1 and extreme bins have low sample sizes as expected. This pattern indicates usable probabilistic outputs already at the inexpensive-feature stage; any minor mid-range bias could be addressed later with lightweight post-hoc calibration if deployment requires tighter probability estimates.

**Fig 5:**
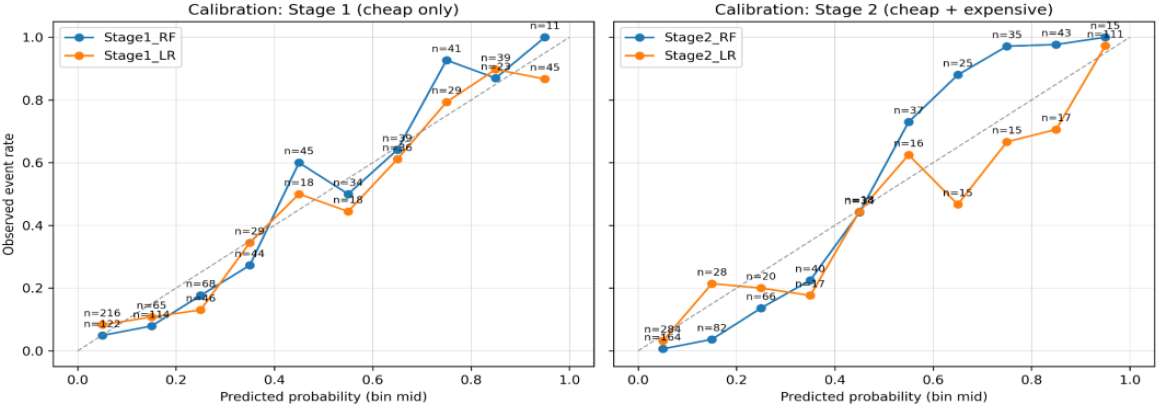
Reliability Diagram Stage 1 (inexpensive only) and Stage 2 (inexpensive + expensive). Observed event rate vs. bin-mid predicted probability; 10 equal-width bins; per-bin n annotated.

In Stage 2 (inexpensive + expensive), discrimination improved and calibration remained strong overall, with a slight over-confidence tendency for Random Forest in the upper-probability bins (>0.7) and a mild under-confidence for Logistic Regression in the mid-high range (≈0.5-0.7) before converging near the diagonal at the top end. These are typical model-specific patterns documented in the calibration literature: tree ensembles can become over-confident and monotonic transformations (e.g., Platt scaling) or non-parametric isotonic regression are standard remedies when strict probability calibration is required [52] (Niculescu-Mizil & Caruana, 2005).

### 3.2 Feature Importance and Selection

We carried out permutation importance analysis for the Stage 1 Random Forest (RF) model using the train-test results (baseline evaluation) over the full set of 24 “inexpensive” predictors ranked Hair growth (Y/N), Skin darkening (Y/N) and Fast food (Y/N) as the most influential features, with mean importance scores of 0.044, 0.031 and 0.020 respectively. In contrast, variables such as Hair loss (Y/N), No. of abortions, Blood group, BP Systolic (mmHg) and Height (cm) were ranked lowest, in some cases contributing negligible or slightly negative mean importance as shown in Fig 6. This ranking highlights a clear separation between high and low-impact predictors, providing an interpretable basis for targeted feature reduction when time or resource constraints limit data collection.

**Fig 6:**
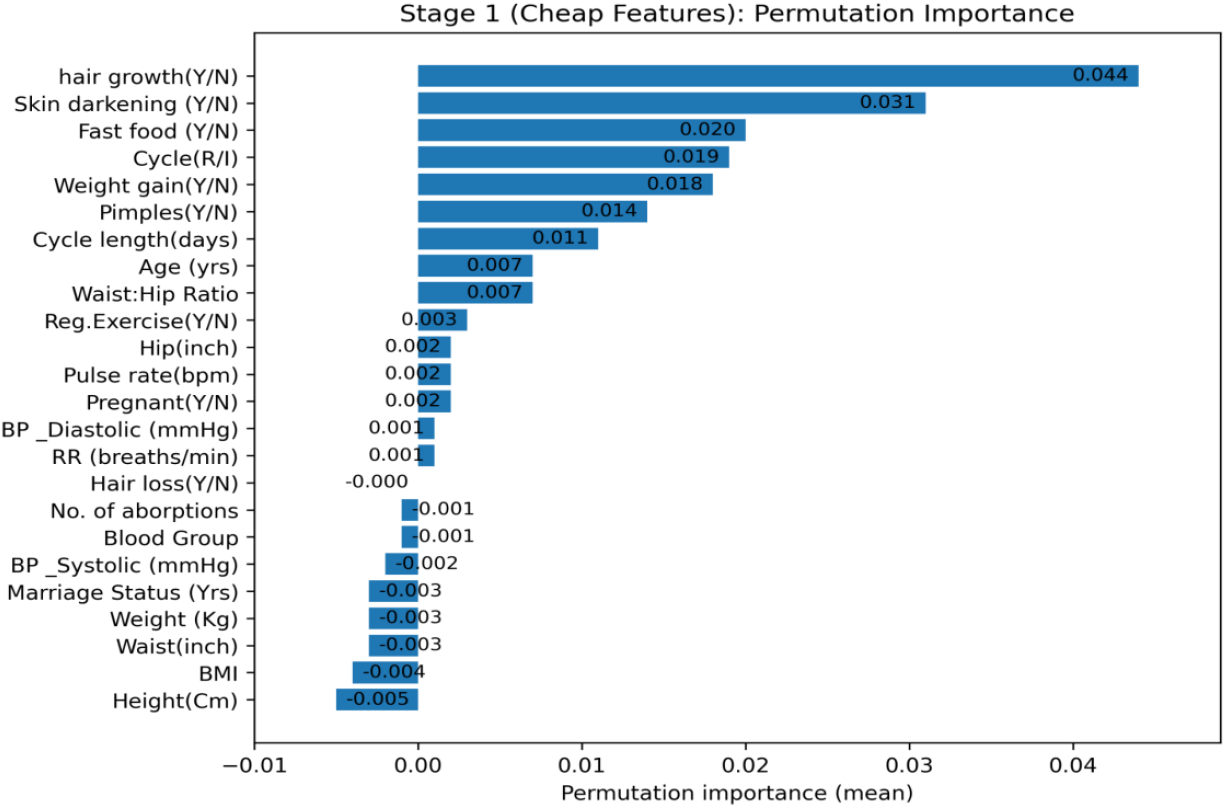
Full ranked list of “inexpensive” features by mean permutation importance (using Stage 1 RF - Baseline Evaluation)

In addition, we conducted a feature drop sensitivity analysis (Fig 7) for Stage 1 RF (Baseline evaluation) showed that removing the bottom 3 to 8 least important features (see Table 9) resulted in no deterioration in discrimination and, in fact, moderate AUC gains: from a baseline of 85.8 to 87.1% (k = 3) and 87.4% (k = 5) while removing the bottom 8 features yielded the highest observed AUC (88.6%) as shown in Fig 8. The associated metric changes revealed small improvements in recall (+7.55% at k = 8) but minor decreases in precision (−4.2% at k = 8) as seen in Fig 7. Brier score and Expected Calibration Error (ECE) changes were negligible across all k values. These results indicate that the least important predictors may introduce noise and their removal can improve model generalisation without sacrificing calibration quality. Notably, body size-related variables such as height, weight, waist and BMI were consistently among the lowest-ranked predictors across the k = 3, 5, and 8 removal settings, indicating substantial redundancy in their contribution to discrimination.

**Table 9:** Least importance features guided by the permutation importance.

| K- removed | Feature dropped |
| --- | --- |
| 3 | Height (cm), BMI, Waist (inch) |
| 5 | Height (cm), BMI, Waist (inch), Weight (kg), Marriage Status (yrs) |
| 8 | Height (cm), BMI, Waist (inch), Weight (kg), Marriage Status (yrs), BP Systolic (mmHg), No. of abortions, Blood Group |

**Fig 7:**
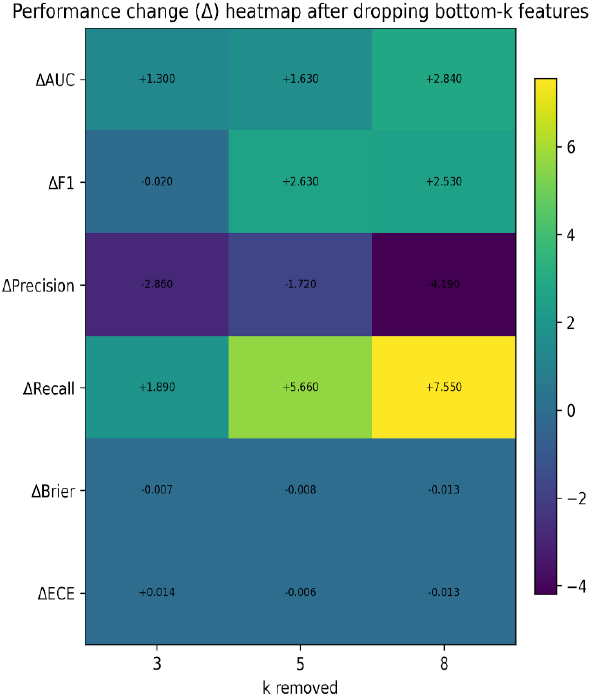
Heatmap of performance metric changes (ΔAUC, ΔF1, ΔPrecision, ΔRecall, ΔBrier, ΔECE) after dropping the k least important features.

**Fig 8:**
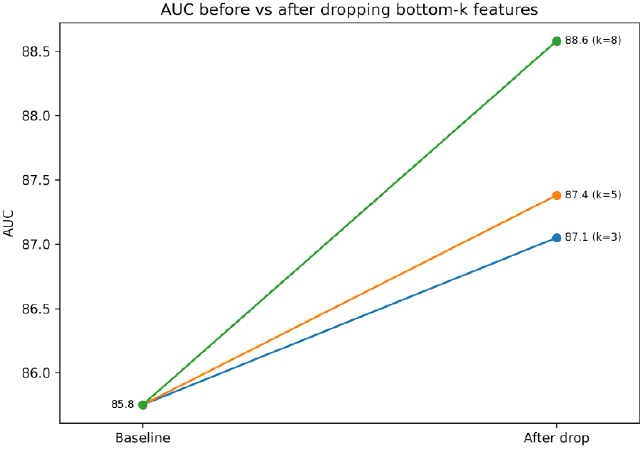
Slope plot of baseline vs. post-drop AUC for k = 3, 5, 8 feature removals.

The AUC slope plot (Fig 8) illustrated a consistent upward trend in performance as low-value predictors were dropped, with diminishing returns beyond the top ~16 features. This supports a modelling approach where reduced feature sets can be deployed at lower operational cost while maintaining, or even slightly enhancing, predictive performance.

To complement the permutation importance and feature drop sensitivity analysis results from Stage 1, we performed an ablation analysis on the Stage 2 Random Forest model, where each “expensive” feature was removed individually to measure its marginal contribution to performance. The change in AUC (ΔAUC) from the full model was used as the sensitivity metric. The results (Table 10) show that Follicle Number (R) had the largest performance drop when removed (ΔAUC = −2.49%), followed by Follicle Number (L) (−1.04%) and PRG (ng/mL) (−0.79%). Several features, including PRL (ng/mL) and Endometrium (mm), had minimal impact (ΔAUC ≤ −0.35%), suggesting they contribute little additional value once stronger predictors are present.

**Table 10:** Ablation results for “expensive” features (using Stage 2 RF - Baseline Evaluation)

| Feature Removed | AUC (%) | $\Delta AUC$ (%) |
| --- | --- | --- |
| Follicle No. (R) | 92.48 | -2.49 |
| Follicle No. (L) | 93.93 | -1.04 |
| PRG (ng/mL) | 94.18 | -0.79 |
| AMH (ng/mL) | 94.19 | -0.78 |
| LH (mIU/mL) | 94.24 | -0.73 |
| FSH/LH | 94.33 | -0.64 |
| II beta-HCG (mIU/mL) | 94.34 | -0.63 |
| RBS (mg/dl) | 94.37 | -0.60 |
| Vit D3 (ng/mL) | 94.37 | -0.60 |
| Avg. F size (R) (mm) | 94.45 | -0.52 |
| TSH (mIU/L) | 94.49 | -0.48 |
| I beta-HCG (mIU/mL) | 94.54 | -0.43 |
| PRL (ng/mL) | 94.63 | -0.34 |
| Hb(g/dl) | 94.64 | -0.33 |
| FSH (mIU/mL) | 94.68 | -0.29 |
| Avg. F size (L) (mm) | 94.74 | -0.23 |
| Endometrium (mm) | 94.83 | -0.14 |

The analysis highlights that a small subset of “expensive” features drives most of the incremental performance gain in Stage 2. Features with negligible ΔAUC could potentially be deprioritised in a cost-sensitive setting without substantial loss in discrimination. These results, when considered alongside permutation importance, provide a consistent view of feature relevance and help prioritise variables for deployment under resource constraints.

### 3.3 Decision Curve Analysis

We carried forward the OOF prediction estimates for the rest of our experimentation and explorations. We designed a decision curve analysis with 2,000 stratified bootstrap resamples and compared the net benefit of Stage 1 and Stage 2 models across threshold probabilities from 0.05 to 0.5. As shown in Fig 9, the decision-curve analysis shows that both models deliver positive clinical net benefit across the examined thresholds (0.05-0.50) compared with the “treat none” strategy (flat at 0). They also outperform “treat all” over nearly the entire range, the green dashed curve declines rapidly with higher thresholds and crosses zero around one-third risk, whereas both RF and LR remain above it. Comparing stages, Stage 2 (inexpensive + expensive) consistently lies above Stage 1 (inexpensive-only), indicating incremental decision value from adding confirmatory tests as seen in Fig 9.

**Fig 9:**
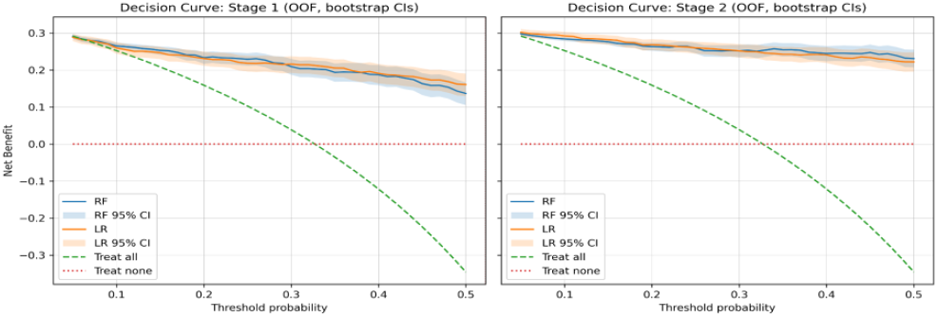
Decision curve plots for Stage 1(left) and Stage 2(right) models with OOF prediction. Net benefit plotted against threshold probability with 95% bootstrap confidence intervals.

The gain is most evident in the 0.15-0.35 region, where many screening decisions are made. At very low thresholds (~0.05-0.10) the curves are close, but Stage 2 still edges higher; at higher thresholds the net benefit naturally tapers for all models, yet Stage 2 maintains the advantage. Between algorithms, the differences are small and the bootstrap CIs overlap across most thresholds. RF is slightly higher than LR in Stage 2, reflecting its stronger precision, while LR tracks closely and sometimes leads in Stage 1, consistent with its higher recall and generally better calibration. Clinically, this suggests a practical strategy: use Stage 1 for low-cost initial screening (it already beats treat-all/none) and when resources allow, escalate to Stage 2 to achieve higher net benefit in the most relevant threshold band.

### 3.4 Capacity-Constrained Triage

We evaluated triage policies under fixed clinic capacity 20%, 40% and 60% of cases that can be advanced using OOF predictions and net benefit (NB) as the objective across thresholds 0.05-0.50 summarised in Table 11. Decision-analytic NB is the right lens here because it directly trades off true positives against false positives at a given risk threshold and avoids accuracy fallacies in imbalanced data [28] Vickers & Elkin, 2006; [53] Vickers et al., 2019)

**Table 11:** Best net benefit by policy and capacity.

| Capacity (% of cohort triaged) | NB (uncertainty aware) | NB (Risk Top-K) | NB (Utility Top-K) | ΔNB (uncertainty aware - Risk Top-K) | ΔNB (uncertainty aware - Utility Top-K) |
| --- | --- | --- | --- | --- | --- |
| 20 | 0.125 | 0.154 | 0.154 | -0.029 | -0.029 |
| 40 | 0.162 | 0.270 | 0.270 | -0.106 | -0.106 |
| 60 | 0.297 | 0.297 | 0.297 | 0.000 | 0.000 |

At all capacities (20%, 40%, 60%), the two baselines (Risk Top-K and Utility Top-K) are essentially indistinguishable: the curves lie on top of each other across thresholds as seen in Fig 10,11 and 12. That matches our definition of the utility score i.e s_i_=p_i_-w(1-p_i_) with a fixed threshold pt: because s_i_ is a monotone transform of pi under symmetric costs, ranking by utility selects the same patients as ranking by risk. The uncertainty-aware policy (prioritising abstentions first, then filling to capacity by utility) underperforms the baselines at every capacity across the shown threshold range (0.05-0.50).

**Fig 10:**
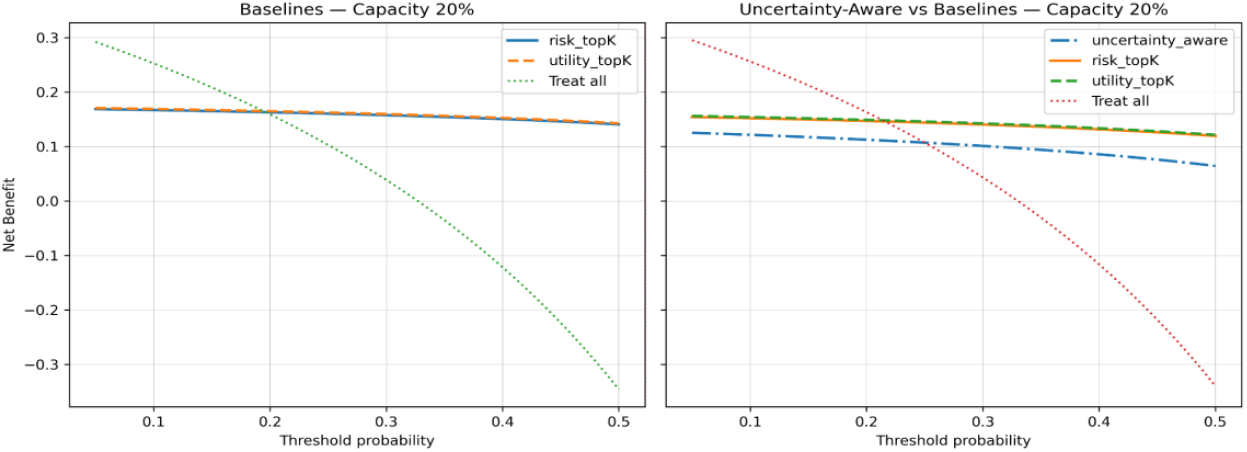
Triage Net Benefit baselines (Risk Top-K vs Utility Top-K) and uncertainty aware, 20% capacity.

**Fig 11:**
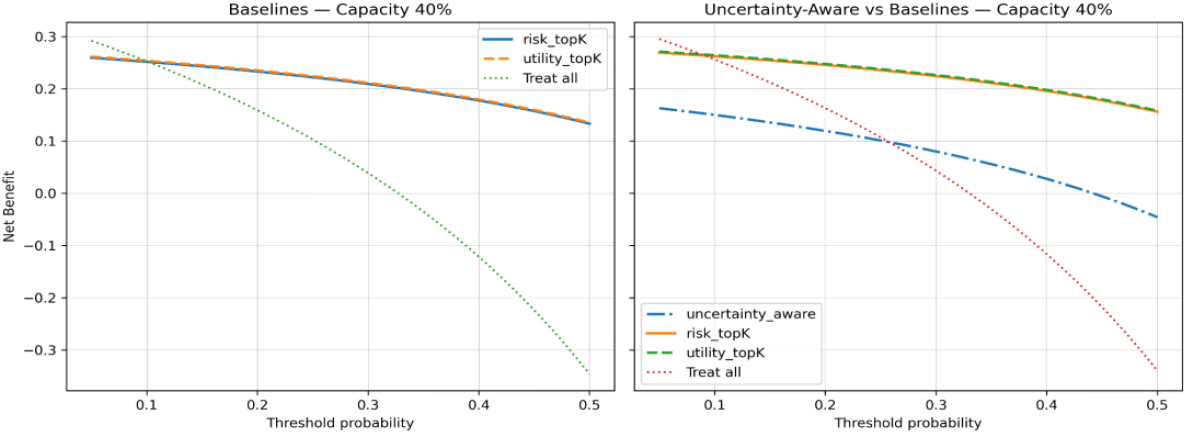
Triage Net Benefit baselines (Risk Top-K vs Utility Top-K) and uncertainty aware, 40% capacity.

**Fig 12:**
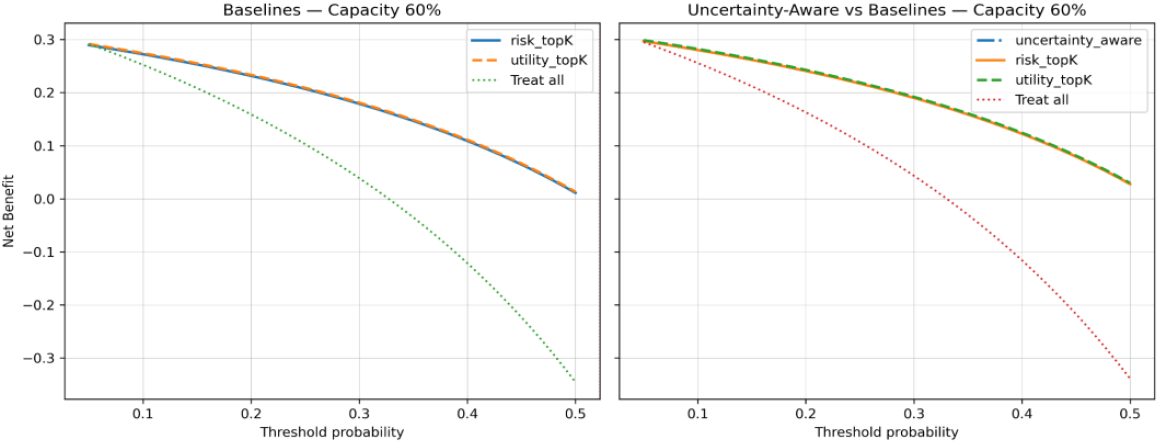
Triage Net Benefit baselines (Risk Top-K vs Utility Top-K) and uncertainty aware, 60% capacity.

At 20% capacity, the Risk Top-K policy dominates the Utility Top-K baseline across thresholds as shown in Fig 10; the dashed utility curve is not visually separable because it overlaps the risk curve. The policy substantially outperforms the “treat all” line. We then compared an uncertainty aware policy i.e. the blue dash-dot curve (penalizing low-confidence cases) to the two baselines. At 20% capacity, uncertainty aware NB is lower than Risk Top-K across most thresholds, suggesting that, when slots are scarce, prioritizing the very highest-risk cases even if some are borderline in confidence yields better NB than discarding them for confidence reasons. At low thresholds the gap is largest; treat-all even exceeds the uncertainty-aware curve until roughly pt≈0.22, demonstrating how costly that diversion is when slots are scarce. This aligns with selective-classification theory: when “coverage” is tight, abstention can reduce true-positive (TP) yield more than it saves false positives (FP) [54] (El-Yaniv, 2010).

At 40% capacity, risk-based selection maintains higher NB than the treat-all reference and the utility curve again lie on top of the risk curve as seen in Fig 11, yielding an indistinguishable trajectory. At 40% capacity, the gap persists: the uncertainty aware curve remains below Risk Top-K for most thresholds, although the margin widens as threshold increases. Prioritising abstentions continues to displace higher-value, higher-risk cases.

At 60% capacity, risk-based selection continues to lead the treat-all line and the utility curve still traces it almost exactly as shown in Fig 12. All three policies converge as evident in Table 11, producing nearly identical NB across thresholds. Once capacity is generous, the top-K sets selected by the different rules overlap strongly and down-weighting uncertain cases has little effect on who gets advanced.

Across panels the net-benefit curves decline monotonically with threshold, so the maximum net benefit occurs at the lowest tested thresholds (~0.05) for each policy/capacity. In this dataset and policy design, rank-by-risk/utility is preferable at constrained capacities, while uncertainty-first selection does not improve and often reduces net benefit; only when capacity is generous do the strategies behave almost identically. These plot-level observations are consistent with the summary results in Table 12, where the optimal threshold was 0.05 for all capacities.

**Table 12:** Best threshold and NB by capacity with True positive and false positive trade-off.

| Capacity | Optimal Threshold | NB@best (Utility Top K) | NB@best (Risk Top K) | TP@best (Utility Top K) | FP@best (Utility Top K) |
| --- | --- | --- | --- | --- | --- |
| 20% | 0.05 | 0.168 | 0.168 | 92 | 16 |
| 40% | 0.05 | 0.259 | 0.259 | 144 | 72 |
| 60% | 0.05 | 0.289 | 0.289 | 165 | 159 |

In our setup, the utility function reduces to sorting by predicted risk with symmetric costs (i.e., the same threshold-induced trade-off used by standard decision curve analysis). Under that symmetry, Utility Top K and Risk Top-K pick the same set of patients at the (empirically optimal) threshold, so their net benefits coincide. This is expected when the policy’s “value” is a monotone transform of risk under fixed class costs and a shared threshold [28] Vickers & Elkin, 2006.

### 3.5 Conformal Prediction: coverage and abstention by age/BMI

We applied class-conditional conformal prediction (α = 0.05) to the Stage 1 Random forest model with OOF predictions (from Table 8) that uses inexpensive features and summarised performance overall and by age and BMI subgroups in Table 13. The conformal predictor achieved a coverage of 94.5% (95% CI: 88.4-98.0%) for the overall count, close to the nominal 95% target. The abstention rate was 41.3% (95% CI: 32.0-51.1%), while the singleton rate was 58.7% (95% CI: 48.9-68.1%), indicating that more than half of predictions were issued as confident single-class outputs while maintaining near-nominal validity.

**Table 13:**
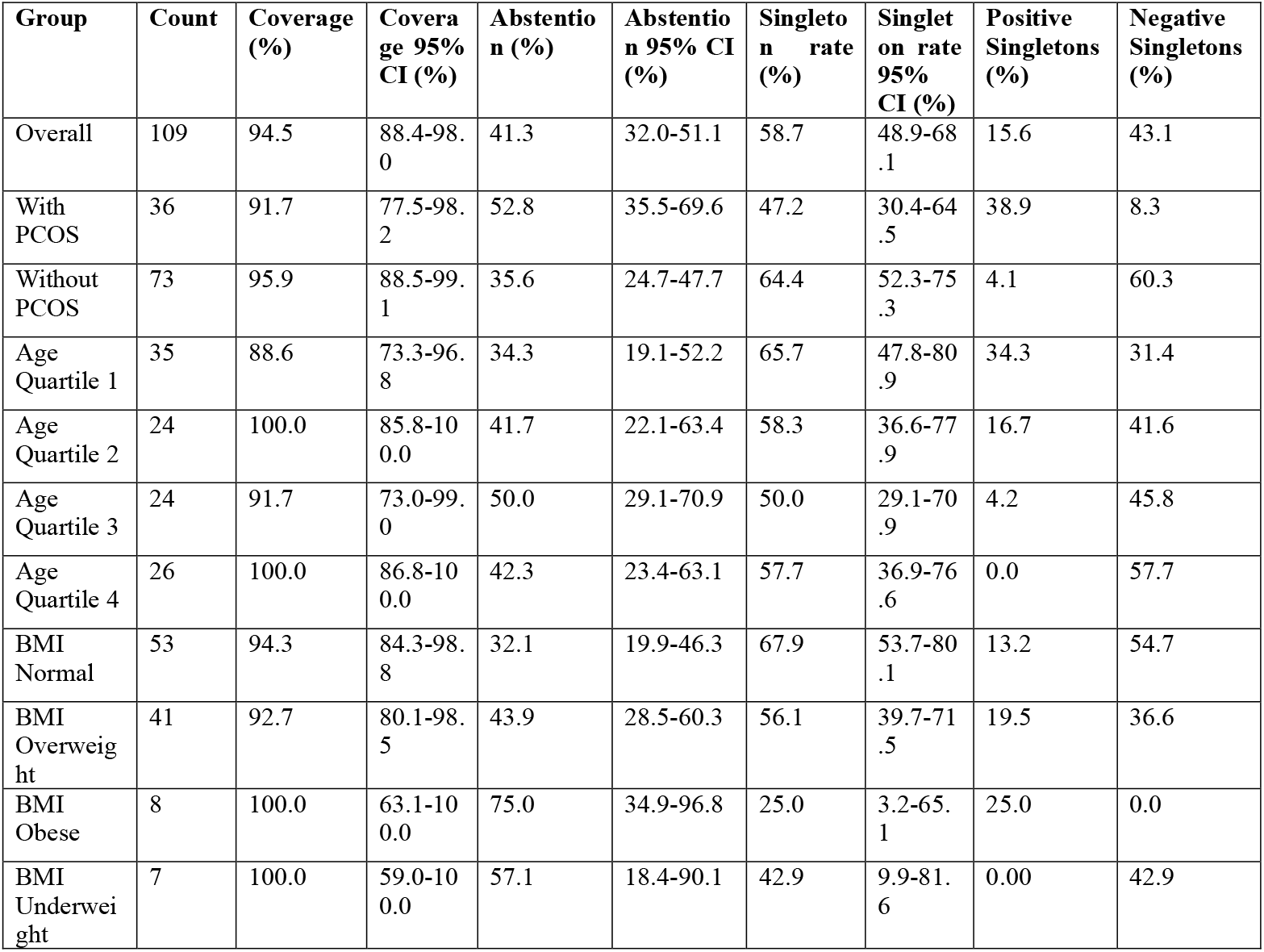
Conformal prediction: overall and subgroup metrics (α=0.05)

Across age quartiles as shown in Fig 13, coverage remained high but varied alongside abstention. Age Q1 achieved 88.6% coverage (95% CI: 73.3-96.8%) with an abstention rate of 34.3%. Age Q2 and Age Q4 both achieved 100% coverage (95% CI: 85.8-100.0% and 86.8-100.0%, respectively), accompanied by abstention rates of 41.7% and 42.3%. Age Q3 recorded 91.7% coverage (95% CI: 73.0-99.0%) with the highest abstention rate at 50.0%, alongside a matched singleton rate of 50.0%. Patterns by BMI subgroup were similar. Participants with normal BMI showed 94.3% coverage (95% CI: 84.3-98.8%), the lowest abstention rate (32.1%), and the highest singleton rate (67.9%). The overweight group achieved 92.7% coverage with an abstention rate of 43.9%. Both obese and underweight groups achieved 100% coverage; however, this was accompanied by substantially higher abstention rates (75.0% and 57.1%, respectively) and wide confidence intervals due to small sample sizes.

**Fig 13:**
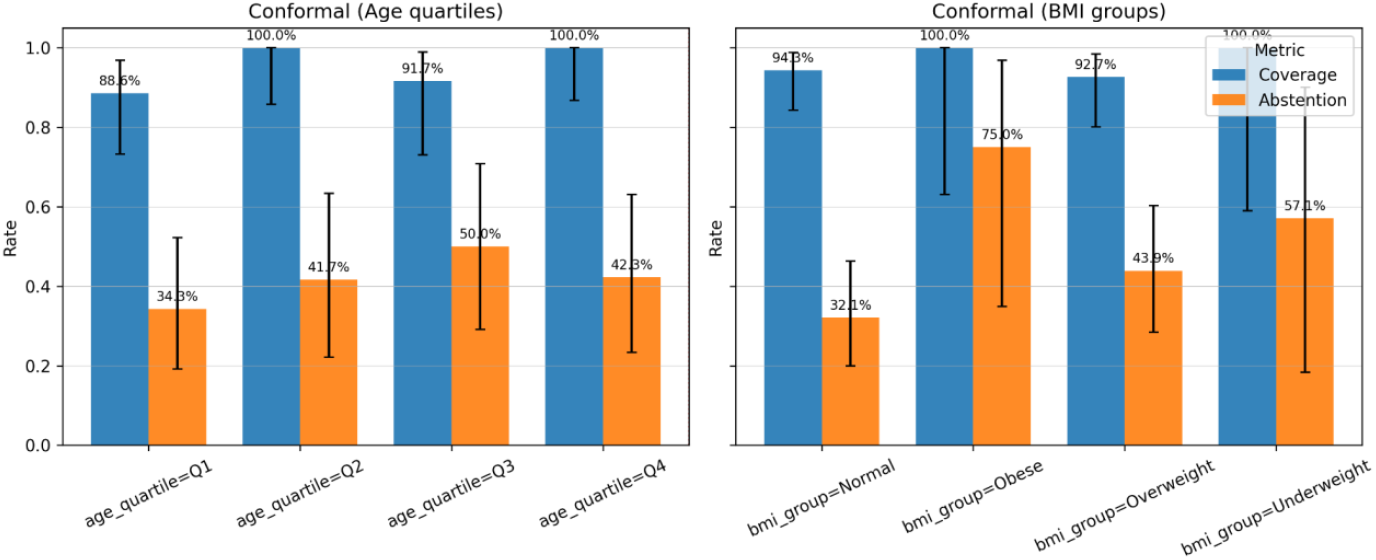
Grouped bars with error bars for coverage and abstention by (left) age quartile and (right) BMI group.

Table 13 further disaggregates singleton predictions into positive and negative assignments and stratifies results by PCOS status. For overall count, 58.7% of predictions were singletons, comprising 15.6% positive singletons and 43.1% negative singletons, indicating that the conformal predictor more frequently issued confident negative predictions at the Stage 1 screening level. Stratification by outcome revealed clear differences. Among individuals with PCOS, abstention increased to 52.8% and the singleton rate fell to 47.2%, with singletons dominated by positive assignments (38.9%) rather than negative ones (8.3%). In contrast, individuals without PCOS exhibited lower abstention (35.6%) and a higher singleton rate (64.4%), with negative singletons predominating (60.3%) and positive singletons occurring infrequently (4.1%).

Across age and BMI subgroups, negative singletons were more common in larger, data-rich strata, whereas smaller or clinically complex subgroups showed higher abstention and fewer confident assignments. This pattern indicates that the conformal prediction mechanism adjusts its decisiveness according to both outcome prevalence and subgroup sample size.

To further characterise the operational behaviour of the conformal prediction, we examined performance conditional on actioned predictions (singleton sets) and abstentions across clinical subgroups. Table 14 summarises abstention counts, the number of actioned cases, selective accuracy on singletons, and error rates among actioned predictions. 64 of 109 cases were actioned as singletons for overall count, with a selective accuracy of 90.6%. Among actioned predictions, false negatives occurred in 17.6% of positive cases, while false positives among negative cases were limited to 6.4%. Across age groups, selective accuracy varied from 82.6% in Age Quartile 1 to 100% in Quartiles 2 and 4. Higher abstention gaps were observed in Age Quartile 3, where selective accuracy dropped to 83.3% and false-negative rates among actioned positives were elevated. Similar patterns were observed across BMI groups, with Normal BMI serving as the lowest-abstention reference. Obese and Underweight subgroups exhibited higher abstention gaps, consistent with small sample sizes and wider uncertainty.

**Table 14:**
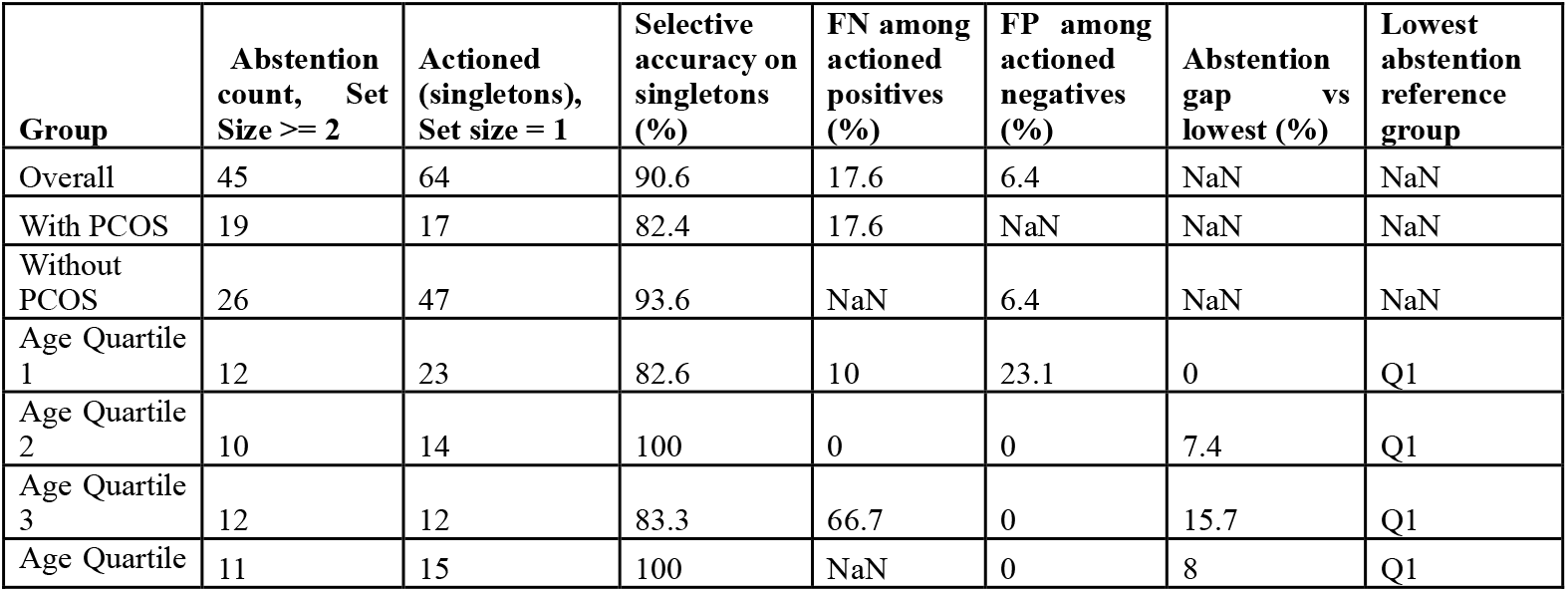
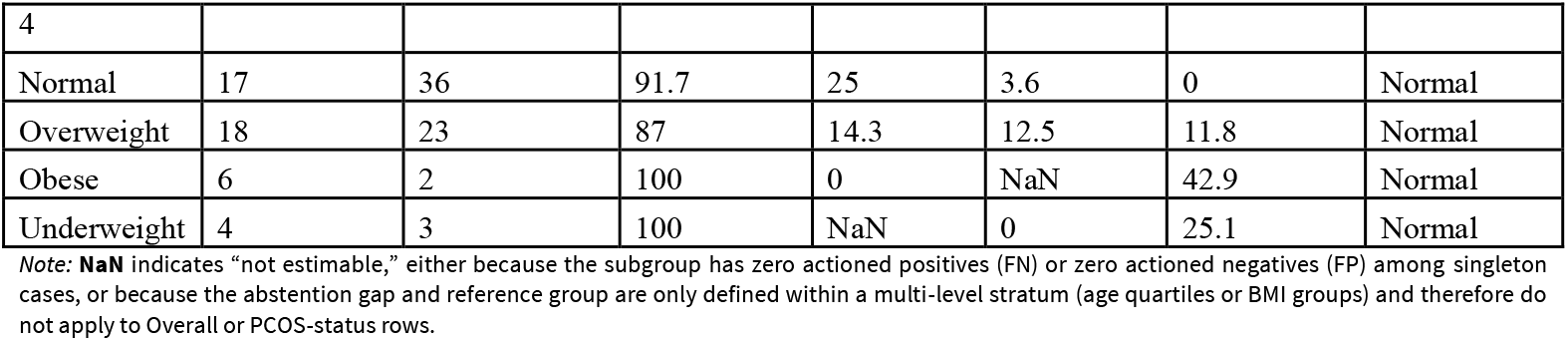
Selective performance metrics under conformal prediction across subgroups.

**Table 15:** Pairwise AUC comparisons using DeLong’s test.

| Model 1 | Model 2 | Model 1 (AUC %) | Model 2 (AUC %) | $\Delta$ AUC (%) | Z Statistic | P value |
| --- | --- | --- | --- | --- | --- | --- |
| Stage 1(RF) | Stage 1(LR) | 88.6 | 87.6 | 1.0 | 0.607 | 0.544 |
| Stage 2(RF) | Stage 2(LR) | 96.0 | 94.5 | 1.5 | 0.965 | 0.334 |
| Stage 2(RF) | Stage 1(RF) | 96.0 | 88.6 | 7.4 | 3.181 | 0.001 |
| Stage 2(LR) | Stage 1(LR) | 94.5 | 87.6 | 6.9 | 2.828 | 0.005 |

To contextualise subgroup effects, we also show the age and BMI distributions by PCOS status (Figs 14-15). The age histogram centres in the late 20s to mid-30s for both classes, while BMI for positives is shifted mildly right; these distributions help explain why coverage is stable but abstention rises in tails where data are sparse.

**Fig 14:**
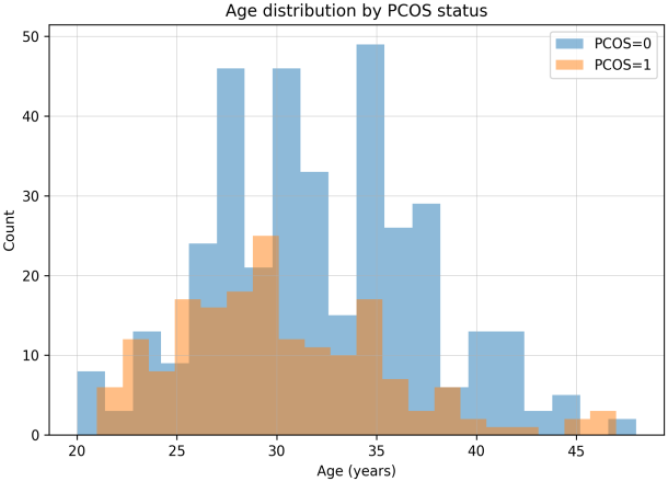
Age distribution by PCOS status.

**Fig 15:**
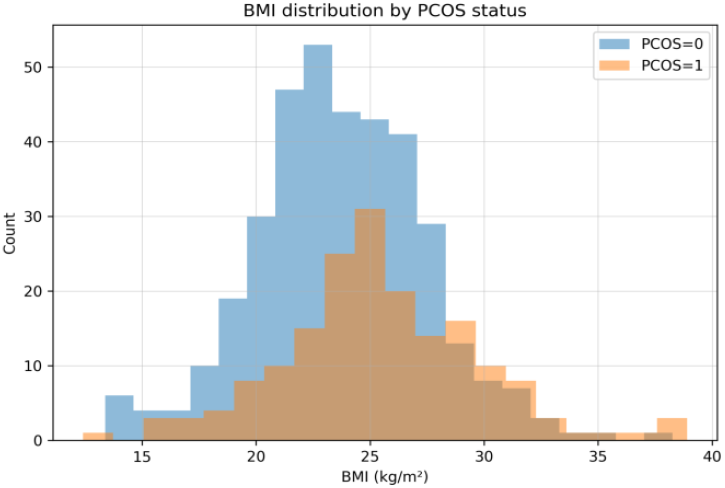
BMI distribution by PCOS status.

Bootstrap resampling showed consistent improvements in Brier scores (Table 16). Stage 2(RF) achieved a −0.041 reduction relative to Stage 1(RF) & Stage 2(LR) a −0.045 reduction compared to Stage 1(LR) indicating more accurate probability estimates. This is consistent with the smaller Expected Calibration Errors (ECE) observed for Stage 2(LR) in Table 17 and supports the more stable decision thresholds identified in DCA.

**Table 16:** Bootstrap differences in Brier scores.

| Model 1 | Model 2 | $\Delta$ Brier (1 - 2) | 95% CI (Low) | 95% CI (High) |
| --- | --- | --- | --- | --- |
| Stage 1(RF) | Stage 1(LR) | 0.003 | -0.006 | 0.011 |
| Stage 2(RF) | Stage 2(LR) | 0.006 | -0.004 | 0.017 |
| Stage 2(RF) | Stage 1(RF) | -0.041 | -0.054 | -0.029 |
| Stage 2(LR) | Stage 1(LR) | -0.045 | -0.062 | -0.027 |

**Table 17:** Calibration scores with 95% confidence intervals.

| Model | Brier | ECE | ECE 95% CI (Low) | ECE 95% CI (High) |
| --- | --- | --- | --- | --- |
| Stage 1(RF) | 0.126 | 0.063 | 0.048 | 0.097 |
| Stage 1(LR) | 0.123 | 0.049 | 0.038 | 0.083 |
| Stage 2(RF) | 0.084 | 0.101 | 0.084 | 0.126 |
| Stage 2(LR) | 0.078 | 0.033 | 0.025 | 0.067 |

### 3.6 Statistical Comparisons

A series of statistical tests were conducted to evaluate whether the observed differences in model performance across metrics and settings were statistically significant. The results are summarised in Tables 15-18.

First, DeLong’s test confirmed that both Stage 2 models significantly outperformed their Stage 1 counterparts in AUC (Table 15). The Stage 2(RF) vs. Stage 1(RF) comparison showed a Δ AUC of 7.4 (p = 0.001), while Stage 2(LR) vs. Stage 1(LR) yielded a Δ AUC of 6.9 (p = 0.005). These gains mirror the upward shifts in the net benefit curves observed in the Decision Curve Analysis (Section 3.3), where Stage 2(RF) consistently dominated across clinically relevant threshold ranges.

Calibration analysis (Table 17) further highlighted Stage 2 (LR)’s superior performance, with the lowest Brier score (0.078) and ECE (0.033). This improved probability calibration explains why Stage 2 models maintained higher net benefits over broader threshold ranges in the DCA curves, reducing over or under-treatment risks.

McNemar’s test (Table 18) supported the classification-level improvements. The Stage 2 (RF) vs Stage 1(RF) comparison revealed a highly significant imbalance in discordant predictions (χ^2^ = 26.882, p<0.001), indicating that the observed NB and AUC improvements were driven by genuine reclassification gains rather than chance.

**Table 18:** McNemar’s test results for Stage 2 vs Stage 1.

| Model 1 | Model 2 | 0→1 Errors | 1→0 Errors | Chi-Square | P- Value |
| --- | --- | --- | --- | --- | --- |
| Stage 1(RF) | Stage 1(LR) | 25 | 12 | 3.892 | 0.143 |
| Stage 2(RF) | Stage 2(LR) | 16 | 21 | 0.432 | 0.806 |
| Stage 2(RF) | Stage 1(RF) | 21 | 72 | 26.882 | 0.000 |
| Stage 2(LR) | Stage 1(LR) | 22 | 55 | 13.299 | 0.001 |

These statistical comparisons corroborate the gains reported in DCA, triage simulations, quantify the magnitude and robustness of integration, reinforcing the conclusion that Stage 2 (RF) provides the most balanced and reliable performance profile for both discrimination and decision support.

## 4.0 Discussion of Result

The staged modelling framework in this study demonstrated that predictive performance can be meaningfully enhanced by moving from a Stage 1 model built solely on low-cost features to a Stage 2 model incorporating richer diagnostics. Across both random forest (RF) and logistic regression (LR) classifiers, Stage 2 models achieved significant AUC gains over Stage 1 (Δ AUC = 7.4% for random forest, p = 0.001; Δ AUC = 6.9% for logistic regression, p = 0.005) as presented in Table 15.

In this work, feature importance and sensitivity analysis revealed that over 80% of Stage 1 predictive performance could be retained with the top 16 inexpensive features freeing administrative time and workload with fewer diagnosis, suggesting potential for cost-sensitive deployment strategies. Removing low-ranked inexpensive features resulted in consistent gains in discrimination. Excluding the bottom three, five, and eight features each produced incremental improvements in AUC (Figure 7 & 8), indicating that several routine variables introduced noise rather than signal. These findings suggest that a reduced set of inexpensive features may yield a more stable and efficient first-line model for PCOS risk assessment, where rapid and accessible screening is clinically advantageous. In addition, correlated body size measures indicated that multicollinearity may reduce their marginal contribution to prediction in this study, even though such variables remain clinically relevant for subgroup definition and downstream analysis.

Similarly, ablation of expensive features produced ΔAUC values ranging from approximately 0.14% to 2.49% as seen in Table 10. A small subset of features produced the largest performance reductions when removed, indicating substantial marginal utility, whereas others produced minimal change and offer limited incremental value. This pattern highlights which costly investigations are most informative for refining PCOS risk estimates and which may be deprioritised when cost or accessibility is a constraint; an important consideration given the variability in diagnostic pathways across clinical settings. These findings support earlier work showing that incremental diagnostic enrichment can boost predictive accuracy without proportionally increasing cost [55] (Desautels T et al., 2016) [56] (Shim T et al 2018). The two stages demonstrate how a structured, cost-aware pipeline can prioritise inexpensive features for early stratification in suspected PCOS cases while reserving high-cost diagnostics for individuals in whom they add demonstrable predictive value. This stepped approach reflects how PCOS assessments are often constrained by resource availability and supports the development of triage strategies that optimise both efficiency and diagnostic yield.

A key observation from this study concerns the behaviour of uncertainty-aware triage under different capacity constraints in the context of PCOS risk assessment. At 20% and 40% capacity (Figure 10 & 11), the uncertainty-aware policy did not outperform risk-based selection; in all threshold regions, its net benefit was lower because abstaining on borderline-confidence cases reduced true-positive yield more than it reduced false positives (Table 11). In a PCOS pathway, this implies that diverting uncertain cases away from immediate assessment may delay identification of individuals with clinically meaningful risk profiles, especially when specialist resources are limited. This behaviour is consistent with selective-classification theory, which predicts that abstention is disadvantageous when coverage is tight. Only at higher capacities (60%), where there is greater overlap between the group of patients selected for further assessment produced by different prioritisation rules, all three policies evaluated in this work converge to similar net benefit and the influence of uncertainty diminish. These findings suggest that uncertainty-aware triage may be most useful in PCOS settings where clinical capacity is sufficient to accommodate abstention without markedly reducing true-positive capture, whereas under more constrained conditions, prioritising high-risk cases remains the more effective operational strategy.

Interestingly, at the 40% capacity level, the model delivers the most informative balance between case yield and over treatment. Net benefit rises by 54% (from 0.168 to 0.259) in Table 12, and when contrasted against the uncertainty aware strategy in Table 11, it produces an incremental gain of 0.106. This operating point captures a substantial increase in true positives relative to the 20% setting, which yielded only 0.029, while maintaining a far more acceptable false positive burden than the 60% configuration, which delivered no incremental gain. In this work, the 40% capacity therefore represents the most clinically efficient region for PCOS treatment allocation, providing meaningful improvement in detected cases without a disproportionate rise in unnecessary interventions.

Building on the triage framework, the conformal prediction analysis further clarifies how uncertainty can be operationalised within the Stage 1 screening layer for PCOS. As shown in Table 13, overall marginal coverage was 94.5%, close to the nominal 95% target, indicating that the Stage 1 model maintained valid uncertainty guarantees despite relying only on low-cost features. At the same time, the abstention rate of 41.3% and singleton rate of 58.7% suggest a meaningful separation between cases that can be actioned immediately and those requiring escalation. This behaviour supports the intended escalation logic; confident singleton predictions can proceed through the low-cost pathway without reliance on costly diagnostics, while abstentions are deferred to Stage 2 for confirmatory testing using higher-cost diagnostics.

The subgroup results reinforce this interpretation while highlighting important equity considerations. Coverage remained high across age and BMI strata, including groups with smaller sample sizes, with several subgroups achieving 100% coverage. However, this was often achieved through increased abstention, particularly in the Obese and Underweight BMI groups, where abstention exceeded 57% and confidence intervals widened due to limited counts. This appropriately reflected increased uncertainty rather than overconfident predictions or model failure, supporting equitable triage and risk handling across heterogeneous patient groups when data are sparse or ambiguous. This ensured that groups with high uncertainty are preferentially escalated rather than forced into potentially unreliable early decisions when low-cost information is insufficient and complements the broader goal of improving the utility of AI-assisted PCOS triage.

Also, the separation of singleton outputs into positive and negative assignments provides additional insight into how uncertainty is handled within the staged framework. At the Stage 1 screening level, confident negative singletons were issued far more frequently than positive ones, particularly among individuals without PCOS. This behaviour is appropriate for a low-cost front-line screen, supporting efficient rule-out while limiting unnecessary escalation.

Furthermore, Table 14 provides insight into how conformal prediction shapes decision quality when predictions are actioned. The high selective accuracy observed overall indicates that restricting decisions to singleton predictions effectively concentrates correctness among cases deemed sufficiently certain by the model. The elevated abstention rate among PCOS-positive individuals suggests that uncertainty-aware screening is conservative in complex cases, prioritising safety by deferring ambiguous presentations to Stage 2 escalation. While this increases abstention, it limits false positives among actioned negatives and supports cautious deployment in diagnostic pathways where missed PCOS cases carry higher clinical cost. Subgroup analysis highlights the trade-off between abstention and reliability. Age and BMI groups with higher abstention gaps also exhibited either reduced selective accuracy or unstable error estimates, demonstrating the role of conformal prediction as a safeguard in data-sparse settings.

A key contribution of this work is demonstrating that targeted, data-driven feature selection can improve discriminatory performance without materially increasing data acquisition cost. Our feature-importance and drop-sensitivity analysis showed that strong predictive performance could be preserved using a reduced set of top-ranked features, supporting the feasibility of accurate yet resource-efficient models in clinical settings where time, cost, and access to diagnostics are constrained. The novelty of this study lies in the unified evaluation of cost-aware feature optimisation, confidence-informed triage, and conformal prediction within a single staged framework. While each of these components has been examined in isolation in prior work, their joint application, empirical assessment and synergy in a capacity-constrained, safety-critical PCOS screening context provides a practical template for deploying uncertainty-aware AI systems that balance efficiency, reliability, and operational constraints.

## 5.0 Limitations & Future Works

This study has several limitations that should be considered when interpreting the findings which reflect the exploratory nature of the study. Rather than providing definitive guidance for deployment, the findings are intended to illustrate the potential of staged, uncertainty-aware AI frameworks as a step toward more efficient, equitable and clinically grounded PCOS diagnostic support.

First, the dataset size was relatively modest (n = 541), which limits statistical power for subgroup analysis, particularly for smaller BMI categories, and constrains the generalisability of the results. External validation on larger and more diverse PCOS cohorts will be essential to assess robustness across populations and care settings. Second, the modelling and operational analysis were conducted on retrospective data. Prospective evaluation will be required to understand how uncertainty-aware triage and staged escalation perform in real clinical workflows, including their effects on diagnostic timing, clinician workload, and patient outcomes. Third, this work deliberately focused on a limited set of well-established algorithms and subgroup definitions to support a proof-of-concept evaluation. Future studies could expand this framework to include additional modelling approaches, comparative analysis across machine learning algorithms at each decision stage, post hoc calibration and alternative uncertainty-handling strategies. Similarly, subgroup analysis could be extended beyond age and BMI to incorporate richer clinical and socio-demographic factors relevant to PCOS heterogeneity and equity considerations.

Furthermore, the PCOS outcome label was derived from the Kaggle dataset, for which detailed clinical diagnostic criteria are not made publicly available/accessible. As a result, label validity cannot be independently verified and may reflect heterogeneous diagnostic standards. This represents a key limitation and motivates future work using clinically curated datasets with explicit diagnostic definitions. In addition, although the framework is designed as a staged pipeline, Stage 2 models were evaluated independently for capacity constrained triaging and decision curve analysis rather than being trained and assessed exclusively on cases escalated from Stage 1. In real-world deployment, escalation would introduce selection effects and partial observability, particularly when high-cost diagnostics are unavailable. Explicit modelling of this selection mechanism remains an important direction for future work. Finally, Decision curve analysis, triage policies, and conformal efficiency are inherently prevalence dependent. The reported results reflect the prevalence structure of the study dataset and may not generalise to settings with different baseline PCOS prevalence. Future work should evaluate prevalence reweighting, recalibration strategies, and scenario-based sensitivity analyses to support deployment in specific clinical contexts.

## 6.0 Conclusion

This study presents a proof-of-concept, cost-efficient AI framework designed to support PCOS risk stratification under operational and capacity constraints. By integrating targeted feature selection, uncertainty-aware triage, and conformal prediction, we demonstrate how predictive performance, reliability, and transparency can be jointly considered within a staged clinical workflow. Rather than introducing new modelling techniques, the framework shows how well-established methods can be structured to prioritise low-cost information, manage uncertainty explicitly, and guide escalation to more resource-intensive assessment when warranted.

Across the staged pipeline, we observed that selective inclusion of inexpensive features improved early discrimination, while a limited set of higher-cost inputs contributed meaningful incremental value at later stages. Decision curve analysis illustrated how triage strategies interact with capacity constraints, and conformal prediction provided near-nominal coverage with interpretable abstention rates across age and BMI subgroups. Together, these components highlight how model evaluation can move beyond accuracy alone to reflect operational feasibility, safety and subgroup stability. While the findings are not intended to support immediate clinical deployment or broad generalisation, they illustrate the potential of structured, uncertainty-aware AI pipelines to inform diagnostic decision-making in resource-constrained settings. In the context of PCOS where diagnostic pathways are often delayed, heterogeneous and costly, this work contributes a practical methodological step toward AI-assisted triage strategies that may ultimately support more timely and equitable assessment in women’s health.

## Data and Code Availability

The datasets analyzed during the current study is publicly available at Kaggle (https://www.kaggle.com/datasets/prasoonkottarathil/polycystic-ovary-syndrome-pcos/data). The code supporting the findings of this study is available from the corresponding author on reasonable request.

## Data Availability

All data produced in the present work are contained in the manuscript

https://www.kaggle.com/datasets/prasoonkottarathil/polycystic-ovary-syndrome-pcos/data

## About The Authors

**Adewale Alex Adegoke** currently works as a Data Systems Manager at the Westminster Foundation for Democracy (WFD) in the UK, where applied his extensive knowledge of Data science and analytics to implement robust data solutions for insights and strategic decision-making that are pivotal to delivering global programmes. Adewale has also worked as a Data Scientist at a UK consulting firm, where he applied advanced Machine learning techniques and research methodologies to develop data-driven solutions for businesses across the world. His research focuses on exploring and examining the reliability and dependability of machine learning systems used in the healthcare and wellbeing domains, with a keen interest in measuring prediction uncertainty, distribution-free confidence methods, and the assessment of model performance. He holds a Master’s degree in Applied Artificial Intelligence and Data Science from Southampton Solent University, which he completed in 2023, as well as a B.Eng. in Civil Engineering from the Federal University of Technology, Minna, Nigeria, in 2011. During his Master’s degree studies, he made significant contributions to several innovative research initiatives, notably in applying natural language processing for emotion detection and sentiment analysis of social media discussions.

**Idris Babalola** is a Senior Data Scientist with the Department of Health and Social Care United Kingdom, where his work focuses on national healthcare data linkage and care pathway analysis. He holds a B.Eng. in Chemical Engineering (2011) from Federal University of Technology Minna Nigeria and an MSc in AI and Data Science from Southampton Solent University UK (2022). He has contributed to EU-funded applied machine learning research with Istya Air, a French-based company focused on indoor air quality (IAQ) prediction. Prototypes developed during this phase were later operationalised and served as a foundation for large-scale deployment during the Olympic Games Paris 2024. He has also previously held part-time academic positions at Southampton Solent University, including MSc Research Supervisor in data science and artificial intelligence, and Associate Lecturer in Computing. His research interests focus on the reliability and trustworthiness of machine learning systems in health, including model uncertainty quantification, conformal prediction. He also works extensively in applied natural language processing, retrieval-augmented generation and API-driven LLM experimentation for real-world healthcare applications. He currently volunteers his service as a peer reviewer for Information Research in Sweden, reviewing AI and data science manuscripts within the information science domain.

**Peter Adebayo Odesola** is a data analyst and applied AI researcher whose work focuses on designing trustworthy, statistically robust machine learning systems for healthcare and other data domains. He holds a BTech in Human Anatomy from Ladoke Akintola University of Technology, Nigeria and a Postgraduate Diploma in Education completed prior to his MSc. in Artificial Intelligence and Data Science from Solent University, Southampton (2022). This combined background in life sciences, education and advanced data science underpins his ability to bridge domain knowledge, methodology and clear scientific communication. His professional experience spans roles as an Independent Researcher in Health, AI and Data Science (in collaboration with data scientists), Data Analyst positions in consultancy and industry, and his current community-focused role with Data Living UK. At Data Living UK, he translates complex open and health-related datasets into actionable insights, designs data dashboards highlighting social and health trends, and supports data storytelling workshops that have reached over 100 learners in underserved communities. Peter regularly collaborates with multidisciplinary teams, including UK civil service data scientists, community organisations and consulting clients, providing analytical leadership from project scoping through to communication of findings.

## Notes

### Competing Interest Statement

The authors have declared no competing interest.

### Funding Statement

This study did not receive any funding

### Summary of Updates

We modified the manuscript title, made clearer areas of the methodology, added a work flow diagram, expanded on the results and revamped the discussion of results.

## References

[1] Lizneva, D., Suturina, L., Walker, W., Brakta, S., Gavrilova-Jordan, L., & Azziz, R. (2016). Criteria, prevalence and phenotypes of polycystic ovary syndrome. Fertility and sterility, 106(1), 6–15. 10.1016/j.fertnstert.2016.05.003

[2] D’Souza, Pramila & Rodrigues Devina & Kaipangala Raja & KC, Leena & Dalmeida, Joylene. (2022). Correlation between the Physical Signs, Clinical Parameters and the Quality of Life in Young Women with Polycystic Ovarian Syndrome: An Explorative Study. Journal of South Asian Federation of Obstetrics and Gynaecology. 14. 17–21. DOI: 10.5005/jp-journals-10006-1993.

[3] Tay, C. T., Garrad, R., Mousa, A., Bahri, M., Joham, A., & Teede, H. (2023). Polycystic ovary syndrome (PCOS): international collaboration to translate evidence and guide future research. Journal of Endocrinology, 257(3), e220232. Retrieved Oct 2, 2025, from 10.1530/JOE-22-0232

[4] Arghavan Ghafari, Malihe Maftoohi, Mohammadamin Eslami Samarin, Sepideh Barani, Majid Banimohammad, Reza Samie. The last update on polycystic ovary syndrome (PCOS), diagnosis criteria and novel treatment, Endocrine and Metabolic Science, Volume 17, (2025), 100228, ISSN 2666-3961, 10.1016/j.endmts.2025.100228.

[5] Long C, Feng H, Duan W, Chen X, Zhao Y, Lan Y, Yue R. Prevalence of polycystic ovary syndrome in patients with type 2 diabetes: A systematic review and meta-analysis. Front Endocrinol (Lausanne). 2022 Aug 31;13:980405. doi: 10.3389/fendo.2022.980405. PMID: 36120432; PMCID: PMC9471325.

[6] Ramadan S. Hussein, Salman Bin Dayel, Othman Abahussein. Polycystic Ovary Syndrome and Reproductive Health: A Comprehensive Review. Clin. Exp. Obstet. Gynecol. 2024, 51(12), 269. DOI: 10.31083/j.ceog5112269

[7] Barrera FJ, Brown EDL, Rojo A, Obeso J, Plata H, Lincango EP, Terry N, Rodríguez-Gutiérrez R, Hall JE and Shekhar S (2023) Application of machine learning and artificial intelligence in the diagnosis and classification of polycystic ovarian syndrome: a systematic review. Front. Endocrinol. 14:1106625. doi: 10.3389/fendo.2023.1106625

[8] Vazquez J, Facelli JC. Conformal Prediction in Clinical Medical Sciences. J Healthc Inform Res. 2022 Jan 28;6(3):241–252. doi: 10.1007/s41666-021-00113-8. PMID: 35898853; PMCID: PMC9309105.

[9] Xu X, Jiang Y, Du J, Sun H, Wang X, Zhang C. Development and validation of a prediction model for suboptimal ovarian response in polycystic ovary syndrome (PCOS) patients undergoing GnRH-antagonist protocol in IVF/ICSI cycles. J Ovarian Res. 2024 May 28;17(1):116. doi: 10.1186/s13048-024-01437-w. PMID: 38807145; PMCID: PMC11134646.

[10] Zad, Z. et al. “Predicting polycystic ovary syndrome with machine learning algorithms from electronic health records.” Frontiers in Endocrinology (2024). 10.3389/fendo.2024.1298628

[11] Suha, S. et al. “Machine learning for early detection of polycystic ovary syndrome using ultrasound image analysis.” Scientific Reports 12, 21817 (2022). 10.1038/s41598-022-21724-0

[12] Abdelsalam, S.H. et al. “Optimized polycystic ovarian disease prognosis and classification using modern AI multi-modality approaches.” BMC Medical Informatics and Decision Making 24, 256 (2024). 10.1186/s12911-024-02577-8

[13] Agirsoy, M.; Oehlschlaeger, M.A. “A machine learning approach for non-invasive PCOS diagnosis from ultrasound and clinical features.” Scientific Reports 15:33638, 2025. DOI: 10.1038/s41598-025-10453-9.

[14] Zigarelli, A.; Jia, Z.; Lee, H. “Machine-Aided Self-diagnostic Prediction Models for Polycystic Ovary Syndrome: Observational Study.” JMIR Formative Research 6(3):e29967 (2022). 10.2196/29967

[15] Shanmugavadivel, K., M S, M.D., T R, M. et al. Optimized polycystic ovarian disease prognosis and classification using AI based computational approaches on multi-modality data. BMC Med Inform Decis Mak 24, 281 (2024). 10.1186/s12911-024-02688-9

[16] Tong, C.; Wu, Y.; Zhuang, Z.; Yu, Y. “A diagnostic model for polycystic ovary syndrome based on machine learning.” Scientific Reports 15:9821, 2025. DOI: 10.1038/s41598-025-92630-4.

[17] Polycystic Ovary Syndrome (PCOS) - Kaggle (Kerala dataset, 541 patients).” Accessed 24th July 2025. https://www.kaggle.com/datasets/prasoonkottarathil/polycystic-ovary-syndrome-pcos/data

[18] Khanna, V.V. et al. “A Distinctive Explainable Machine Learning Framework for Detection of Polycystic Ovary Syndrome.” Applied System Innovation 6(2):32 (2023). 10.3390/asi6020032

[19] Rahman, M.M. et al. “Empowering Early Detection: A Web-Based Machine Learning Approach for PCOS Prediction.” Informatics in Medicine Unlocked 47:101500 (2024). 10.1016/j.imu.2024.101500

[20] Taha, R.; Zain El Abdin, H.; Musleh, T. “Comparative Analysis of Supervised Machine Learning Models for PCOS Prediction Using Clinical Data.” Journal of Engineering Research and Sciences 4(6):16–26, 2025. DOI: 10.55708/js0406003.

[21] Tiwari, S. et al. “SPOSDS: A Smart Polycystic Ovary Syndrome Diagnostic System Using Machine Learning.” Expert Systems with Applications 203:117592 (2022). 10.1016/j.eswa.2022.117592

[22] Danaei Mehr, H.; Polat, H. “Diagnosis of polycystic ovary syndrome through different machine learning and feature selection techniques.” Health and Technology 12, 137–150 10.1007/s12553-021-00613-y (2022).

[23] Bharati, S.; Podder, P.; Mondal, M.R.H. “Diagnosis of Polycystic Ovary Syndrome Using Machine Learning Algorithms.” 2020 IEEE Region 10 Symposium (TENSYMP) (2020). 10.1109/TENSYMP50017.2020.9230932

[24] Mohi Uddin K.M.; Bhuiyan, M.T.A.; Rahman, M.M.; Islam, M.M.; Uddin, M.A. “Early PCOS Detection: A Comparative Analysis of Traditional and Ensemble Machine Learning Models With Advanced Feature Selection.” Engineering Reports 7(2):e70008, 2025. DOI: 10.1002/eng2.70008.

[25] Panjwani, B.; Yadav, J.; Mohan, V.; Agarwal, N.; Agarwal, S. “Optimized Machine Learning for the Early Detection of Polycystic Ovary Syndrome in Women.” Sensors 25(4):1166, 2025. DOI: 10.3390/s25041166.

[26] Emara, H.M.; El-Shafai, W.; Soliman, N.F.; Algarni, A.D.; Alkanhel, R.; Abd El-Samie, F.E. “A stacked learning framework for accurate classification of polycystic ovary syndrome with advanced data balancing and feature selection techniques.” Frontiers in Physiology 16:1435036, 2025. DOI: 10.3389/fphys.2025.1435036.

[27] Angelopoulos, A.N.; Bates, S. “Conformal Prediction: A Gentle Introduction.” Foundations and Trends in Machine Learning 16(4):494–591 (2022). 10.1561/2200000101

[28] Vickers, A.J.; Elkin, E.B. “Decision Curve Analysis: A Novel Method for Evaluating Prediction Models.” Medical Decision Making 26(6):565–574 (2006). 10.1177/0272989X06295361

[29] Van Calster Ben & Mclernon David & van Smeden Maarten & Wynants Laure & Steyerberg, Ewout. (2019). Calibration: The Achilles heel of predictive analytics. BMC Medicine. 17. DOI:10.1186/s12916-019-1466-7.

[30] Moons KG, Altman DG, Reitsma JB, Ioannidis JP, Macaskill P, Steyerberg EW, Vickers AJ, Ransohoff DF, Collins GS. Transparent Reporting of a multivariable prediction model for Individual Prognosis or Diagnosis (TRIPOD): explanation and elaboration. Ann Intern Med. 2015 Jan 6;162(1):W1–73. doi: 10.7326/M14-0698. PMID: 25560730.

[31] Steyerberg, E. W. (2019). Clinical Prediction Models: A Practical Approach to Development, Validation and Updating (2nd ed.). Springer. DOI: 10.1007/978-3-030-16399-0.

[32] Moons KG, Altman DG, Vergouwe Y, Royston P. Prognosis and prognostic research: application and impact of prognostic models in clinical practice. BMJ. 2009 Jun 4;338:b606. doi: 10.1136/bmj.b606. PMID: 19502216.

[33] Olsson, H., Kartasalo, K., Mulliqi, N. et al. Estimating diagnostic uncertainty in artificial intelligence assisted pathology using conformal prediction. Nat Commun 13, 7761 (2022). 10.1038/s41467-022-34945-8

[34] Zhang Z. Missing data imputation: focusing on single imputation. Ann Transl Med. 2016 Jan;4(1):9. doi: 10.3978/j.issn.2305-5839.2015.12.38. PMID: 26855945; PMCID: PMC4716933.

[35] Breiman, L. (2001). Random forests. Machine Learning, 45(1), 5–32. 10.1023/A:1010933404324

[36] Powers, David & Ailab,. (2011). Evaluation: From precision, recall and F-measure to ROC, informedness, markedness & correlation. J. Mach. Learn. Technol. 2. 2229–3981. DOI 10.9735/2229-3981.

[37] Brier, G. W. (1950). Verification of forecasts expressed in terms of probability. Monthly Weather Review, 78(1), 1–3

[38] Naeini, M. P., Cooper, G. F., & Hauskrecht, M. (2015). Obtaining well calibrated probabilities using Bayesian binning. AAAI Conference on Artificial Intelligence.

[39] Altmann, A., Tolosi, L., Sander, O., & Lengauer, T. (2010). Permutation importance: a corrected feature importance measure. Bioinformatics, 26(10), 1340–1347. 10.1093/bioinformatics/btq134

[40] Kaufman, S., Rosset, S., Perlich, C., & Stitelman, O. (2012). Leakage in data mining: Formulation, detection and avoidance. ACM Transactions on Knowledge Discovery from Data, 6(4), 1–21.

[41] Vovk, V., Gammerman, A., & Shafer, G. (2022). Algorithmic Learning in a Random World. Springer Science & Business Media. DOI:10.1007/978-3-031-06649-8

[42] Shafer, G., & Vovk, V. (2008). A tutorial on conformal prediction. Journal of Machine Learning Research, 9(Mar), 371–421.

[43] Rina Foygel Barber, Emmanuel J. Candès, Aaditya Ramdas, Ryan J. Tibshirani “Predictive inference with the jackknife+,” The Annals of Statistics, Ann. Statist. 49(1), 486–507, (February 2021)

[44] Clopper, C. J., & Pearson, E. S. (1934). The use of confidence or fiducial limits illustrated in the case of the binomial. Biometrika, 26(4), 404–413.

[45] DeLong, E. R., DeLong, D. M., & Clarke-Pearson, D. L. (1988). Comparing the areas under two or more correlated receiver operating characteristic curves: A nonparametric approach. Biometrics, 44(3), 837–845.

[46] Wasserstein, R. L., Schirm, A. L., & Lazar, N. A. (2019). Moving to a World Beyond “p < 0.05”. The American Statistician, 73(up1), 1–19.

[47] Collins GS, Reitsma JB, Altman DG, Moons KG. Transparent reporting of a multivariable prediction model for individual prognosis or diagnosis (TRIPOD): the TRIPOD statement. BMJ. 2015 Jan 7;350:g7594. doi: 10.1136/bmj.g7594. PMID: 25569120.

[48] Tom Fawcett, An introduction to ROC analysis, Pattern Recognition Letters, Volume 27, Issue 8, 2006, Pages 861–874, ISSN 0167-8655, 10.1016/j.patrec.2005.10.010.

[49] Noroozi, Z., Orooji, A., & Erfannia, L. (2023). Analyzing the impact of feature selection methods on machine learning algorithms for heart disease prediction. Scientific Reports, 13, 22588. 10.1038/s41598-023-49962-w

[50] Cutler, D. R., Edwards, T. C., Jr, Beard K.H., Cutler, A., Hess, K. T., Gibson, J., & Lawler, J. J. (2007). Random forests for classification in ecology. Ecology, 88(11), 2783–2792. 10.1890/07-0539.1

[51] García-Carretero, R., Holgado-Cuadrado, R., & Barquero-Pérez Ó. (2021). Assessment of Classification Models and Relevant Features on Nonalcoholic Steatohepatitis Using Random Forest. Entropy, 23(6), 763. 10.3390/e23060763

[52] Alexandru Niculescu-Mizil and Rich Caruana. 2005. Predicting good probabilities with supervised learning. In Proceedings of the 22nd international conference on Machine learning (ICML ‘05). Association for Computing Machinery, New York, NY, USA, 625–632. DOI:10.1145/1102351.1102430

[53] Vickers, A.J., Van Calster, B. & Steyerberg, E.W. A simple, step-by-step guide to interpreting decision curve analysis. Diagn Progn Res 3, 18 (2019). 10.1186/s41512-019-0064-7

[54] El-Yaniv, R., & Wiener, Y. (2010). On the Foundations of Noise-free Selective Classification. J. Mach. Learn. Res., 11, 1605–1641.

[55] Desautels T, Calvert J, Hoffman J, Jay M, Kerem Y, Shieh L, Shimabukuro D, Chettipally U, Feldman MD, Barton C, Wales DJ, Das R. Prediction of Sepsis in the Intensive Care Unit With Minimal Electronic Health Record Data: A Machine Learning Approach. JMIR Med Inform. 2016 Sep 30;4(3):e28. doi: 10.2196/medinform.5909. PMID: 27694098; PMCID: PMC5065680.

[56] Shim H., Hwang S.J., Yang E., 2018. “Joint Active Feature Acquisition and Classification with Variable-Size Set Encoding.” 32nd Conference on Neural Information Processing Systems (NeurIPS 2018), Montréal, Canada

